# Health benefits of a five-day at-home modified fasting program: a randomised controlled trial

**DOI:** 10.1101/2024.11.01.24316348

**Authors:** Franziska Grundler, Quinten R. Ducarmon, Alfred Holley, Marie Knufinke, Selina Strathmeyer, Steffen Heelemann, Roland Geyer, Borja Martinez Tellez, Michael R MacArthur, Georg Zeller, Françoise Wilhelmi de Toledo, Robin Mesnage

**Affiliations:** Buchinger Wilhelmi Clinic, Wilhelm-Beck-Straße 27, 88662 Überlingen, Germany; Structural and Computational Biology Unit, European Molecular Biology Laboratory, Heidelberg, Germany; Leiden University Center for Infectious Diseases (LUCID), Leiden University Medical Center, Leiden, the Netherlands; Laboratory of Nutrition and Metabolic Epigenetics, Department of Health Sciences and Technology, ETH Zurich, Zurich, Switzerland; Lifespin GmbH, Am BioPark 13, 93053 Regensburg, Germany; Department of Nursing, Physiotherapy and Medicine and SPORT Research Group (CTS-1024), CIBIS Research Center, University of Almería, Almería, Spain; Department of Medicine, Division of Endocrinology and Einthoven Laboratory for Experimental Vascular Medicine, Leiden University Medical Center, Leiden, The Netherlands; Lewis-Sigler Institute for Integrative Genomics, Princeton University, Princeton NJ 08540, USA; Department of Nutritional Sciences, School of Life Course Sciences, Faculty of Life Sciences and Medicine, King’s College London, SE1 9NH London, UK

**Keywords:** fasting, cardiometabolic health, FMD, gut microbiome, weight loss, metabolomics

## Abstract

**Background:** Fasting has been shown to be one of the most cost-effective methods to improve cardiometabolic health. We studied a 5-day hypocaloric (∼600 kcal/day) and ketogenic, modified fasting program (MFP) for at-home interventions. We hypothesised that this MFP induces metabolic changes comparable to 5 days of prolonged fasting (75-250kcal/day).

**Methods:** We tested the MFP in a two-arm randomised controlled trial where sixty-four healthy subjects were randomised to MFP or control group. Serum biochemistry analyses and questionnaires allowed for determining effects on cardiometabolic risk factors. Emotional well-being, possible side effects and physical activity were assessed with questionnaires. Biological pathways and metabolic processes were explored with nuclear magnetic resonance blood metabolomics and gut metagenomics analyses.

**Results:** MFP participants (n=32) experienced weight loss (−3.1 ± 1.1 kg) persisting one month after the intervention. The MFP induced significant, but transient reductions, in systolic (−6.4 ± 11.5 mmHg) and diastolic (−4.4 ± 7.7 mmHg) blood pressure, glucose levels, HbA1c and coagulation factors. The MFP led to greater reductions in BMI (p=0.006), diastolic pressure (p = 0.009), cholesterol (p = 0.02), and LDL (p = 0.02) in individuals at risk of cardiometabolic diseases compared to healthy individuals. Total cholesterol, LDL-C and HDL-C levels continued decreasing during food reintroduction. The MFP also increased emotional and physical well-being. Blood metabolomics revealed a significant decrease in chronic inflammation markers. Shotgun metagenomics of the gut microbiome showed trends in the changes in relative abundance of the majority of bacterial species and their genomic repertoire of carbohydrate-active enzymes (CAZymes). This reflected a decrease in families metabolising dietary fibre substrates and an increase in families metabolising host-derived glycan substrates. Comparing MFP effects with a previous cohort’s 5-day prolonged fasting showed similar metabolic changes.

**Conclusion:** This MFP is safe and effectively improves cardiometabolic health and emotional well-being in healthy individuals. It offers comparable metabolic benefits to those observed during 5-day prolonged fasting in a clinic. It is safe to be practised at home, widely accessible and compatible with individuals’ everyday life.

## Introduction

Throughout history, humans and free living animals have regularly undergone long periods of fasting in response to limited food availability, during several days or weeks according to seasonal cycles ^1^, and several hours while they sleep in alignment with circadian rhythms ^2^. In current times, the disruption in life rhythmicity brought by the western lifestyle and the occurrence of “chronic fasting deficiency”— when individuals do not respect physiologic fasting periods (e.g. eating at night or skipping longer circannual fasting periods)—can be hypothesised to contribute to the development of cardiometabolic diseases, along with other factors such as poor nutrition, exposure to toxic pollutants, decreased sleep duration and sedentarism ^3^. Conversely, calorie restriction and fasting are known to extend lifespan and health span in various animal models ^1,4^.

Fasting strategies vary widely in terms of duration and frequency, as well as in caloric intake and macronutrient profiles ^5^. These can range from daily time-restricted eating, which reduces eating windows thus allowing fasting periods each day ^6,7^, to extended fasting periods lasting up to 21 days or more ^8,9^. Retaining the health benefits of fasting is often difficult once subjects return to their adipogenic environment ^10^. Incorporating fasting strategies in everyday life under online supervision could improve compliance and increase the range of applications of fasting interventions and thus promote healthy longevity in the general population ^11,12^.

We previously reported the safety, well-being and metabolic improvements in a cohort of 1422 patients undergoing prolonged fasting programme (PF) with the Buchinger Wilhelmi protocol (75-250 kcal daily as fruit juice, vegetable soup and honey) lasting between 4 and 21 days under medical supervision and moderate physical activity ^8^. Whether these effects persist after fasting—like after any intensive nutritional intervention—depends largely on the diet during the period of food reintroduction and the adherence to a healthy lifestyle post-intervention ^13^. This underscores the necessity for new, easily accessible interventions that can be routinely implemented in daily life. To address this need, a five-day, 600-calorie ketogenic diet with restricted protein and carbohydrate intake, was designed to mimic a 5-day period of PF programme in an at-home setting ^8^.

Mechanisms of fasting-induced health benefits are well understood. At the molecular level, the restriction of glucose and proteins causes the inactivation of nutrient-sensing pathways, which in return modulate adaptive cellular responses such as DNA repair, autophagy, apoptosis, and oxidative stress defence ^14^. These mechanisms are recognized hallmarks of ageing which can partially explain the healthy longevity-enhancing effects of fasting ^15^. In addition, the switch from food-derived glucose to endogenous energy substrates, mainly fatty acids, triggers the production of ketone bodies which, beyond energy provision, signals fasting adaptations, reducing oxidative stress and inflammation ^4^. The ability of the body to switch between different fuel sources, such as carbohydrates and fats, is known as metabolic flexibility and can be trained by regular fasting bouts ^16,17^.

Our two-arm randomised controlled trial (RCT) aimed to explore the effects of this MFP on cardiometabolic risk factors, well-being, gut microbiome composition and functional potential as well as the feasibility of this MFP at home. In order to understand whether the effects of our MFP can induce hallmarks of fasting metabolism, we performed blood biochemistry, blood metabolomics and a gut metagenomics analysis.

## RESULTS

### Study participation

A total of 82 healthy subjects were screened for eligibility between March 2023 and August 2023. Sixteen subjects were excluded due to limited availability during the trial (n=10), drug intake (n=4), recent antibiotic use (n=1), and pregnancy (n=1). Two subjects declined participation due to loss of motivation and an acute cold. We randomly assigned 64 participants to a group receiving the MFP or to a group of participants who were told to continue with their usual eating behaviour and lifestyle (control group). Recruitment, randomization and follow-up participation are summarised in the Consolidated Standards of Reporting Trials (CONSORT) diagram (Figure 1A) together with a timeline of the programme phases (Figure 1B).

**Figure 1.**
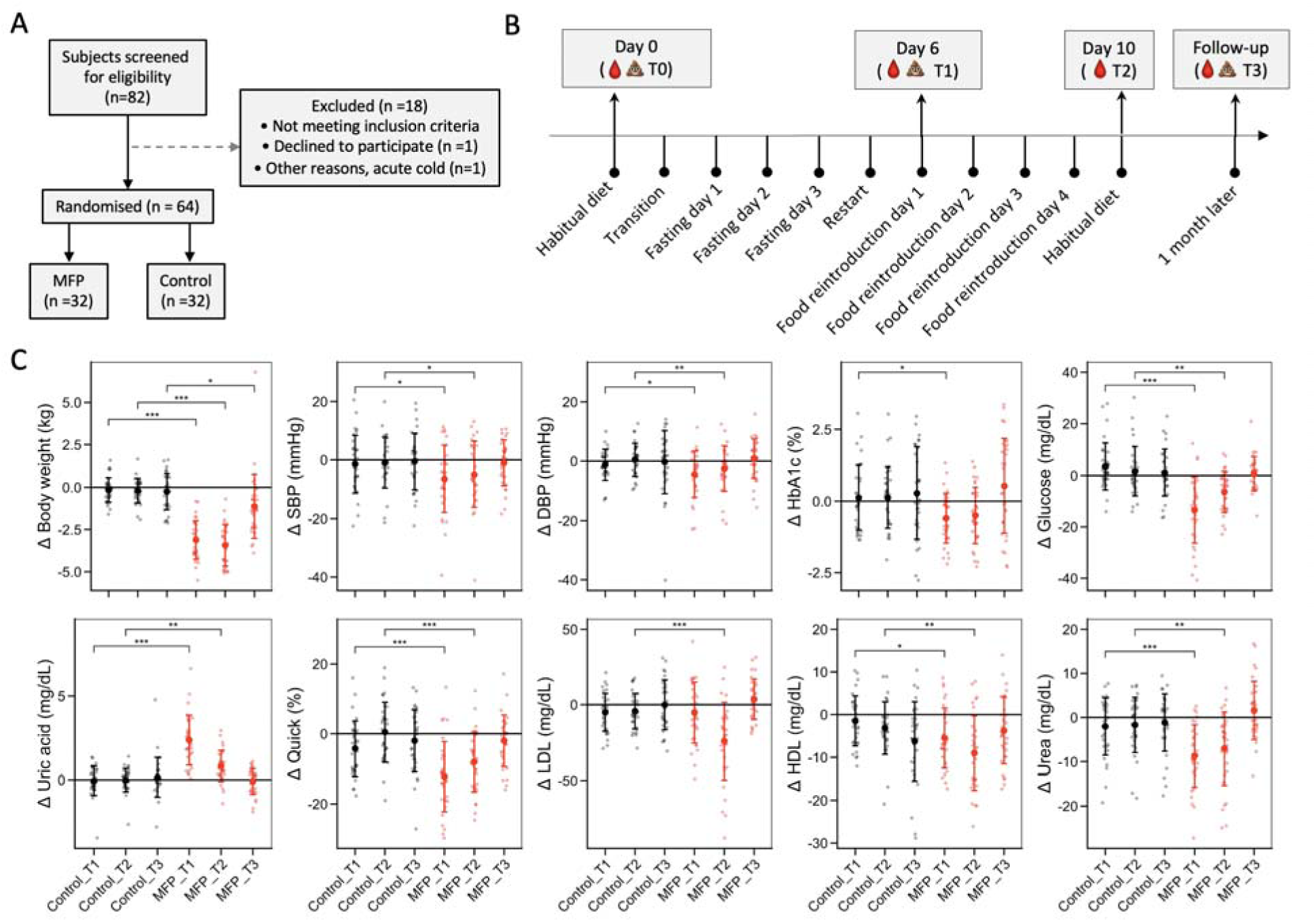
Metabolic effects of the MFP. **A.** Diagram of the recruitment procedure. **B.** Study design with repeated sampling of blood and faecal samples. **C.** Effects of the MFP (red) in comparison to the control group (black) at the end of the MFP period (T1), after 5 days food reintroduction (T2), or a month later (T3), for major parameters reflecting the switch to the fasting metabolism, presented as changes in the mean ± SD together with individual differences for each study individual relative to baseline (T0). The p-value indicates the statistical significance in a linear-mixed model where the effects are compared to the control group at a given timepoint (*, p < 0.05 ; **, p < 0.01 ; ***, p < 0.001).

The 22 men (34.4%) and 42 women (65.6%) who participated in this RCT were mostly Caucasian (62 out of 64), non-smokers and moderate alcohol consumers (Table S4). There were no significant differences in baseline characteristics between the MFP and control group with respect to age, sex, body weight, blood pressure and waist circumference (Table S4). Only BMI was slightly higher in the MFP group than in the control group (Table 1).

**Table 1.**
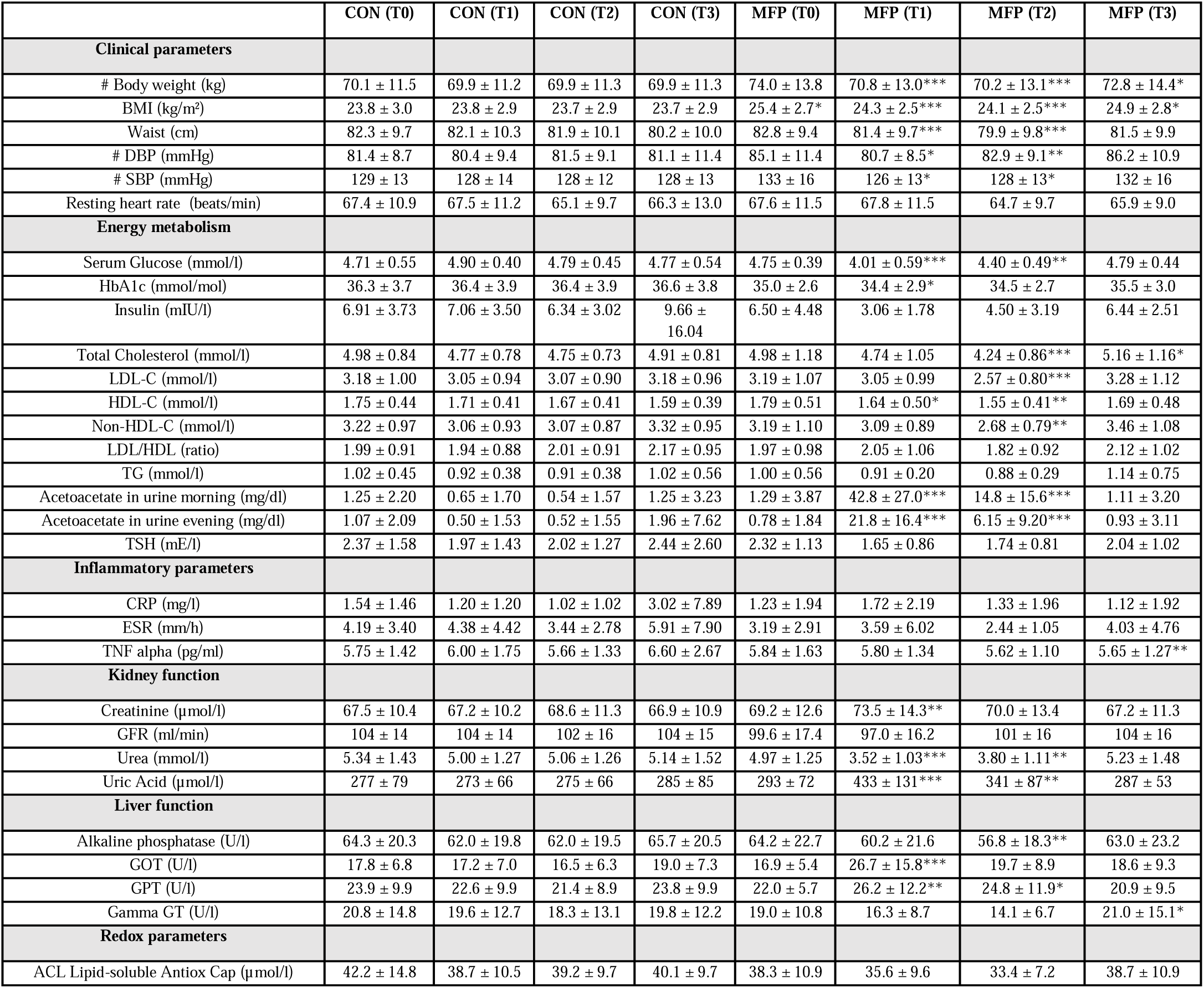
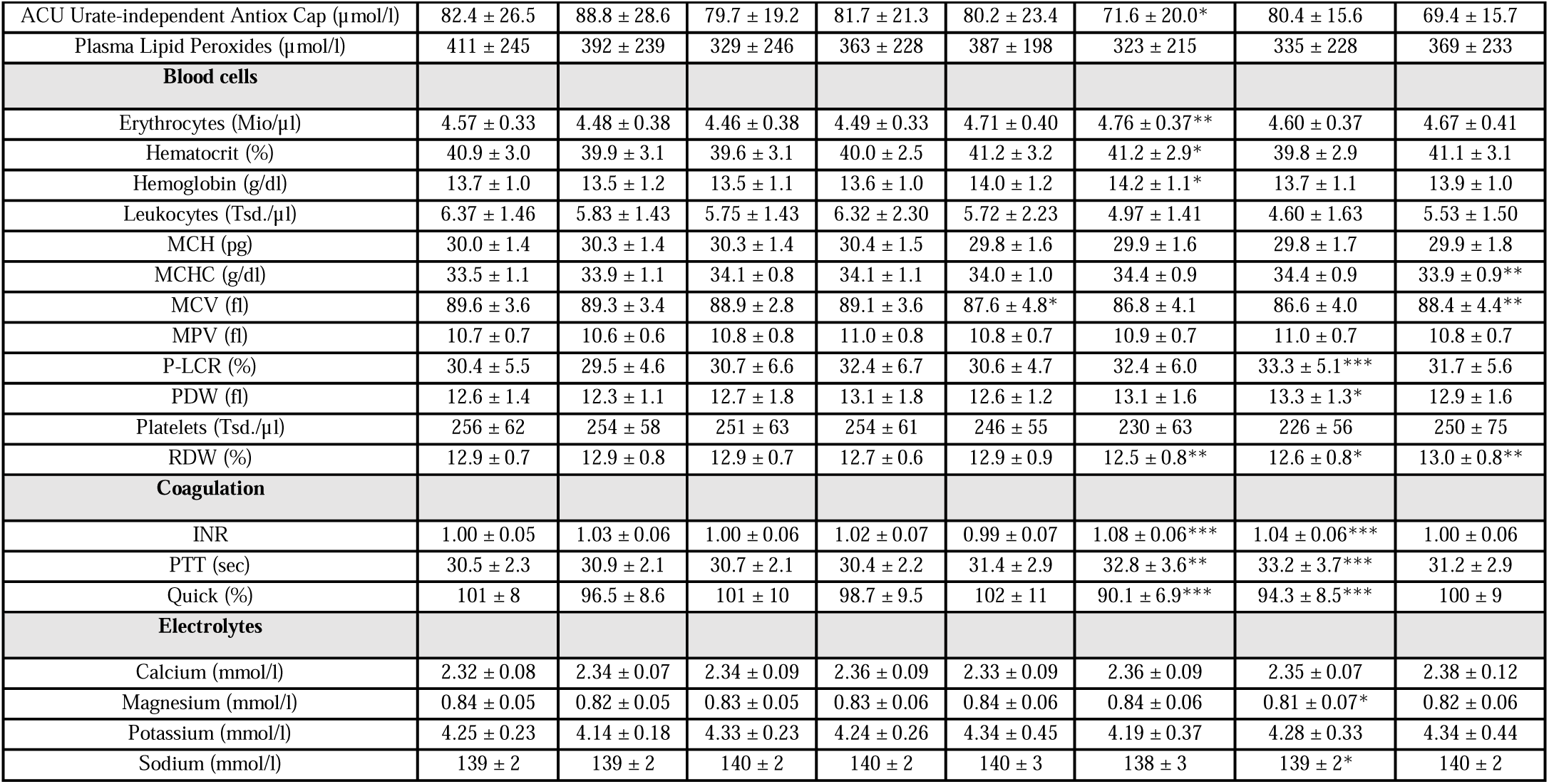
Biomarker or risk factor changes in the study participants. The p-value indicates the statistical significance in a linear-mixed model (individuals as random effects) where the effects of the MFP are compared to the control (CON) group at the same timepoint (*, p < 0.05 ; **, p < 0.01 ; ***, p < 0.001). Data as mean ± SD. # Primary endpoints.

|  | CON (T0) | CON (T1) | CON (T2) | CON (T3) | MFP (T0) | MFP (T1) | MFP (T2) | MFP (T3) |
| --- | --- | --- | --- | --- | --- | --- | --- | --- |
| **Clinical parameters** |  |  |  |  |  |  |  |  |
| # Body weight (kg) | 70.1 ± 11.5 | 69.9 ± 11.2 | 69.9 ± 11.3 | 69.9 ± 11.3 | 74.0 ± 13.8 | 70.8 ± 13.0*** | 70.2 ± 13.1*** | 72.8 ± 14.4* |
| BMI (kg/m<sup>2</sup>) | 23.8 ± 3.0 | 23.8 ± 2.9 | 23.7 ± 2.9 | 23.7 ± 2.9 | 25.4 ± 2.7* | 24.3 ± 2.5*** | 24.1 ± 2.5*** | 24.9 ± 2.8* |
| Waist (cm) | 82.3 ± 9.7 | 82.1 ± 10.3 | 81.9 ± 10.1 | 80.2 ± 10.0 | 82.8 ± 9.4 | 81.4 ± 9.7*** | 79.9 ± 9.8*** | 81.5 ± 9.9 |
| # DBP (mmHg) | 81.4 ± 8.7 | 80.4 ± 9.4 | 81.5 ± 9.1 | 81.1 ± 11.4 | 85.1 ± 11.4 | 80.7 ± 8.5* | 82.9 ± 9.1** | 86.2 ± 10.9 |
| # SBP (mmHg) | 129 ± 13 | 128 ± 14 | 128 ± 12 | 128 ± 13 | 133 ± 16 | 126 ± 13* | 128 ± 13* | 132 ± 16 |
| Resting heart rate (beats/min) | 67.4 ± 10.9 | 67.5 ± 11.2 | 65.1 ± 9.7 | 66.3 ± 13.0 | 67.6 ± 11.5 | 67.8 ± 11.5 | 64.7 ± 9.7 | 65.9 ± 9.0 |
| **Energy metabolism** |  |  |  |  |  |  |  |  |
| Serum Glucose (mmol/l) | 4.71 ± 0.55 | 4.90 ± 0.40 | 4.79 ± 0.45 | 4.77 ± 0.54 | 4.75 ± 0.39 | 4.01 ± 0.59*** | 4.40 ± 0.49** | 4.79 ± 0.44 |
| HbA1c (mmol/mol) | 36.3 ± 3.7 | 36.4 ± 3.9 | 36.4 ± 3.9 | 36.6 ± 3.8 | 35.0 ± 2.6 | 34.4 ± 2.9* | 34.5 ± 2.7 | 35.5 ± 3.0 |
| Insulin (mIU/l) | 6.91 ± 3.73 | 7.06 ± 3.50 | 6.34 ± 3.02 | 9.66 ± 16.04 | 6.50 ± 4.48 | 3.06 ± 1.78 | 4.50 ± 3.19 | 6.44 ± 2.51 |
| Total Cholesterol (mmol/l) | 4.98 ± 0.84 | 4.77 ± 0.78 | 4.75 ± 0.73 | 4.91 ± 0.81 | 4.98 ± 1.18 | 4.74 ± 1.05 | 4.24 ± 0.86*** | 5.16 ± 1.16* |
| LDL-C (mmol/l) | 3.18 ± 1.00 | 3.05 ± 0.94 | 3.07 ± 0.90 | 3.18 ± 0.96 | 3.19 ± 1.07 | 3.05 ± 0.99 | 2.57 ± 0.80*** | 3.28 ± 1.12 |
| HDL-C (mmol/l) | 1.75 ± 0.44 | 1.71 ± 0.41 | 1.67 ± 0.41 | 1.59 ± 0.39 | 1.79 ± 0.51 | 1.64 ± 0.50* | 1.55 ± 0.41** | 1.69 ± 0.48 |
| Non-HDL-C (mmol/l) | 3.22 ± 0.97 | 3.06 ± 0.93 | 3.07 ± 0.87 | 3.32 ± 0.95 | 3.19 ± 1.10 | 3.09 ± 0.89 | 2.68 ± 0.79** | 3.46 ± 1.08 |
| LDL/HDL (ratio) | 1.99 ± 0.91 | 1.94 ± 0.88 | 2.01 ± 0.91 | 2.17 ± 0.95 | 1.97 ± 0.98 | 2.05 ± 1.06 | 1.82 ± 0.92 | 2.12 ± 1.02 |
| TG (mmol/l) | 1.02 ± 0.45 | 0.92 ± 0.38 | 0.91 ± 0.38 | 1.02 ± 0.56 | 1.00 ± 0.56 | 0.91 ± 0.20 | 0.88 ± 0.29 | 1.14 ± 0.75 |
| Acetoacetate in urine morning (mg/dl) | 1.25 ± 2.20 | 0.65 ± 1.70 | 0.54 ± 1.57 | 1.25 ± 3.23 | 1.29 ± 3.87 | 42.8 ± 27.0*** | 14.8 ± 15.6*** | 1.11 ± 3.20 |
| Acetoacetate in urine evening (mg/dl) | 1.07 ± 2.09 | 0.50 ± 1.53 | 0.52 ± 1.55 | 1.96 ± 7.62 | 0.78 ± 1.84 | 21.8 ± 16.4*** | 6.15 ± 9.20*** | 0.93 ± 3.11 |
| TSH (mE/l) | 2.37 ± 1.58 | 1.97 ± 1.43 | 2.02 ± 1.27 | 2.44 ± 2.60 | 2.32 ± 1.13 | 1.65 ± 0.86 | 1.74 ± 0.81 | 2.04 ± 1.02 |
| **Inflammatory parameters** |  |  |  |  |  |  |  |  |
| CRP (mg/l) | 1.54 ± 1.46 | 1.20 ± 1.20 | 1.02 ± 1.02 | 3.02 ± 7.89 | 1.23 ± 1.94 | 1.72 ± 2.19 | 1.33 ± 1.96 | 1.12 ± 1.92 |
| ESR (mm/h) | 4.19 ± 3.40 | 4.38 ± 4.42 | 3.44 ± 2.78 | 5.91 ± 7.90 | 3.19 ± 2.91 | 3.59 ± 6.02 | 2.44 ± 1.05 | 4.03 ± 4.76 |
| TNF alpha (pg/ml) | 5.75 ± 1.42 | 6.00 ± 1.75 | 5.66 ± 1.33 | 6.60 ± 2.67 | 5.84 ± 1.63 | 5.80 ± 1.34 | 5.62 ± 1.10 | 5.65 ± 1.27** |
| **Kidney function** |  |  |  |  |  |  |  |  |
| Creatinine (μmol/l) | 67.5 ± 10.4 | 67.2 ± 10.2 | 68.6 ± 11.3 | 66.9 ± 10.9 | 69.2 ± 12.6 | 73.5 ± 14.3** | 70.0 ± 13.4 | 67.2 ± 11.3 |
| GFR (ml/min) | 104 ± 14 | 104 ± 14 | 102 ± 16 | 104 ± 15 | 99.6 ± 17.4 | 97.0 ± 16.2 | 101 ± 16 | 104 ± 16 |
| Urea (mmol/l) | 5.34 ± 1.43 | 5.00 ± 1.27 | 5.06 ± 1.26 | 5.14 ± 1.52 | 4.97 ± 1.25 | 3.52 ± 1.03*** | 3.80 ± 1.11** | 5.23 ± 1.48 |
| Uric Acid (μmol/l) | 277 ± 79 | 273 ± 66 | 275 ± 66 | 285 ± 85 | 293 ± 72 | 433 ± 131*** | 341 ± 87** | 287 ± 53 |
| **Liver function** |  |  |  |  |  |  |  |  |
| Alkaline phosphatase (U/l) | 64.3 ± 20.3 | 62.0 ± 19.8 | 62.0 ± 19.5 | 65.7 ± 20.5 | 64.2 ± 22.7 | 60.2 ± 21.6 | 56.8 ± 18.3** | 63.0 ± 23.2 |
| GOT (U/l) | 17.8 ± 6.8 | 17.2 ± 7.0 | 16.5 ± 6.3 | 19.0 ± 7.3 | 16.9 ± 5.4 | 26.7 ± 15.8*** | 19.7 ± 8.9 | 18.6 ± 9.3 |
| GPT (U/l) | 23.9 ± 9.9 | 22.6 ± 9.9 | 21.4 ± 8.9 | 23.8 ± 9.9 | 22.0 ± 5.7 | 26.2 ± 12.2** | 24.8 ± 11.9* | 20.9 ± 9.5 |
| Gamma GT (U/l) | 20.8 ± 14.8 | 19.6 ± 12.7 | 18.3 ± 13.1 | 19.8 ± 12.2 | 19.0 ± 10.8 | 16.3 ± 8.7 | 14.1 ± 6.7 | 21.0 ± 15.1* |
| **Redox parameters** |  |  |  |  |  |  |  |  |
| ACL Lipid-soluble Antiox Cap (μmol/l) | 42.2 ± 14.8 | 38.7 ± 10.5 | 39.2 ± 9.7 | 40.1 ± 9.7 | 38.3 ± 10.9 | 35.6 ± 9.6 | 33.4 ± 7.2 | 38.7 ± 10.9 |
| ACU Urate-independent Antiox Cap (μmol/l) | 82.4 ± 26.5 | 88.8 ± 28.6 | 79.7 ± 19.2 | 81.7 ± 21.3 | 80.2 ± 23.4 | 71.6 ± 20.0* | 80.4 ± 15.6 | 69.4 ± 15.7 |
| Plasma Lipid Peroxides (μmol/l) | 411 ± 245 | 392 ± 239 | 329 ± 246 | 363 ± 228 | 387 ± 198 | 323 ± 215 | 335 ± 228 | 369 ± 233 |
| **Blood cells** |  |  |  |  |  |  |  |  |
| Erythrocytes (Mio/μl) | 4.57 ± 0.33 | 4.48 ± 0.38 | 4.46 ± 0.38 | 4.49 ± 0.33 | 4.71 ± 0.40 | 4.76 ± 0.37** | 4.60 ± 0.37 | 4.67 ± 0.41 |
| Hematocrit (%) | 40.9 ± 3.0 | 39.9 ± 3.1 | 39.6 ± 3.1 | 40.0 ± 2.5 | 41.2 ± 3.2 | 41.2 ± 2.9* | 39.8 ± 2.9 | 41.1 ± 3.1 |
| Hemoglobin (g/dl) | 13.7 ± 1.0 | 13.5 ± 1.2 | 13.5 ± 1.1 | 13.6 ± 1.0 | 14.0 ± 1.2 | 14.2 ± 1.1* | 13.7 ± 1.1 | 13.9 ± 1.0 |
| Leukocytes (Tsd./μl) | 6.37 ± 1.46 | 5.83 ± 1.43 | 5.75 ± 1.43 | 6.32 ± 2.30 | 5.72 ± 2.23 | 4.97 ± 1.41 | 4.60 ± 1.63 | 5.53 ± 1.50 |
| MCH (pg) | 30.0 ± 1.4 | 30.3 ± 1.4 | 30.3 ± 1.4 | 30.4 ± 1.5 | 29.8 ± 1.6 | 29.9 ± 1.6 | 29.8 ± 1.7 | 29.9 ± 1.8 |
| MCHC (g/dl) | 33.5 ± 1.1 | 33.9 ± 1.1 | 34.1 ± 0.8 | 34.1 ± 1.1 | 34.0 ± 1.0 | 34.4 ± 0.9 | 34.4 ± 0.9 | 33.9 ± 0.9** |
| MCV (fl) | 89.6 ± 3.6 | 89.3 ± 3.4 | 88.9 ± 2.8 | 89.1 ± 3.6 | 87.6 ± 4.8* | 86.8 ± 4.1 | 86.6 ± 4.0 | 88.4 ± 4.4** |
| MPV (fl) | 10.7 ± 0.7 | 10.6 ± 0.6 | 10.8 ± 0.8 | 11.0 ± 0.8 | 10.8 ± 0.7 | 10.9 ± 0.7 | 11.0 ± 0.7 | 10.8 ± 0.7 |
| P-LCR (%) | 30.4 ± 5.5 | 29.5 ± 4.6 | 30.7 ± 6.6 | 32.4 ± 6.7 | 30.6 ± 4.7 | 32.4 ± 6.0 | 33.3 ± 5.1*** | 31.7 ± 5.6 |
| PDW (fl) | 12.6 ± 1.4 | 12.3 ± 1.1 | 12.7 ± 1.8 | 13.1 ± 1.8 | 12.6 ± 1.2 | 13.1 ± 1.6 | 13.3 ± 1.3* | 12.9 ± 1.6 |
| Platelets (Tsd./μl) | 256 ± 62 | 254 ± 58 | 251 ± 63 | 254 ± 61 | 246 ± 55 | 230 ± 63 | 226 ± 56 | 250 ± 75 |
| RDW (%) | 12.9 ± 0.7 | 12.9 ± 0.8 | 12.9 ± 0.7 | 12.7 ± 0.6 | 12.9 ± 0.9 | 12.5 ± 0.8** | 12.6 ± 0.8* | 13.0 ± 0.8** |
| **Coagulation** |  |  |  |  |  |  |  |  |
| INR | 1.00 ± 0.05 | 1.03 ± 0.06 | 1.00 ± 0.06 | 1.02 ± 0.07 | 0.99 ± 0.07 | 1.08 ± 0.06*** | 1.04 ± 0.06*** | 1.00 ± 0.06 |
| PTT (sec) | 30.5 ± 2.3 | 30.9 ± 2.1 | 30.7 ± 2.1 | 30.4 ± 2.2 | 31.4 ± 2.9 | 32.8 ± 3.6** | 33.2 ± 3.7*** | 31.2 ± 2.9 |
| Quick (%) | 101 ± 8 | 96.5 ± 8.6 | 101 ± 10 | 98.7 ± 9.5 | 102 ± 11 | 90.1 ± 6.9*** | 94.3 ± 8.5*** | 100 ± 9 |
| **Electrolytes** |  |  |  |  |  |  |  |  |
| Calcium (mmol/l) | 2.32 ± 0.08 | 2.34 ± 0.07 | 2.34 ± 0.09 | 2.36 ± 0.09 | 2.33 ± 0.09 | 2.36 ± 0.09 | 2.35 ± 0.07 | 2.38 ± 0.12 |
| Magnesium (mmol/l) | 0.84 ± 0.05 | 0.82 ± 0.05 | 0.83 ± 0.05 | 0.83 ± 0.06 | 0.84 ± 0.06 | 0.84 ± 0.06 | 0.81 ± 0.07* | 0.82 ± 0.06 |
| Potassium (mmol/l) | 4.25 ± 0.23 | 4.14 ± 0.18 | 4.33 ± 0.23 | 4.24 ± 0.26 | 4.34 ± 0.45 | 4.19 ± 0.37 | 4.28 ± 0.33 | 4.34 ± 0.44 |
| Sodium (mmol/l) | 139 ± 2 | 139 ± 2 | 140 ± 2 | 140 ± 2 | 140 ± 3 | 138 ± 3 | 139 ± 2* | 140 ± 2 |

### Adverse events

We observed transient adverse events. A woman experienced brief nausea and vomiting on the second day of the MFP. Three participants working together in the same workplace developed symptoms on day 2 of the study, including a woman (control group) who reported nausea, vomiting, headache, dizziness and increased body temperature, a woman (MFP group) who reported dizziness, and a man (MFP group) who reported dizziness and nausea. The latter person took medication (metamizol dipyrone 500 mg for 3 days). All continued their participation. An environmental cause for these adverse events cannot be excluded, especially given that symptoms occurred in one participant of the control group.

### Improved metabolic health in MFP as compared to controls

Participants in the MFP group experienced a variety of improvements in cardiometabolic health which were not observed in the control group (Table 1). Body weight was decreased by 3.1 ± 1.1 kg (p =5.8×10^−87^) by MFP, while systolic and diastolic blood pressure were reduced by 6.4 ± 11.5 mmHg (p=0.03) and 4.4 ± 7.7 mmHg (p=0.05), respectively (Figure 1D and Table 1). Ketone bodies measured in urine with ketone test stripes showed that participants in the MFP group entered ketosis rapidly from day 2 onwards (Figure S1). Blood glucose and HbA1c significantly decreased by 0.71 ± 0.69 mmol/l (p=3.7×10^−57^) and 0.58 ± 0.89 mmol/mol (p=5.5×10^−7^), respectively (Table 1), reflecting the known improvement of glycaemic control caused by fasting.

The participants exhibited characteristic features of fasting metabolism, such as an increase in uric acid levels (p=6.6×10^−18^), while ketonuria (p=1.7×10^−29^) and creatinine slightly increased (p=0.001) (Table 1). There was also an increase in international normalised ratio (INR) (p=9.6×10^−7^) and partial thromboplastin time (PTT) (p=0.001), which altogether reflected mild anticoagulative effects characteristic of the fasting metabolism. The increase in uric acid levels is another hallmark of fasting metabolism which has been associated with an increased antioxidative capacity and reduced lipid peroxidation in previous fasting studies ^18^.

This MFP programme also includes a food reintroduction phase (Fig 1B), which consists of a stepwise reintroduction of different food items, gradually increasing the number of calories (800 to 1600 kcal per day during 4 days). Most of the effects observed at the end of the MFP were stable or continuously improving after the 4 days of food reintroduction (Table 1), including further reductions in body weight, BMI and waist circumference. LDL-C (p=2.2×10^−5^) and non-HDL-C (p=0.002) levels were significantly reduced compared to baseline after food reintroduction (Table 1), while they were only non-significantly reduced at the end of the MFP, suggesting that the MFP can have lipid metabolism-normalising effects detectable mainly after food reintroduction.

To explore the effects of two food reintroduction strategies after fasting, participants were evenly divided: half received a ketogenic diet, and the other half received a non-ketogenic diet (Table S2). Limited differences were observed between the ketogenic and standard diet groups (Table S5). Both groups continued to excrete urinary ketone bodies consistently. Notably, the increase in PTT seen during the modified fasting program (MFP) persisted and was amplified after ketogenic food reintroduction but not with the standard diet (p=0.003). Conversely, the decrease in urea levels observed during MFP was further amplified with standard food reintroduction but not with the ketogenic diet (p=0.0001). Altogether, these findings can suggest that reintroducing food with a ketogenic diet can prolong the health benefits of fasting by sustaining ketosis.

We investigated if the MFP-induced effects persisted after a month (Table 1). Body weight and BMI of the participants remained lower than baseline. Blood pressure returned to pre-intervention levels. Most of the parameters in these healthy subjects, which reflected beneficial changes in metabolism caused by the MFP intervention, returned to baseline levels.

### Effects on individuals at risk of chronic diseases

We evaluated whether the MFP caused even larger benefits for the health of individuals with risk factors of cardiometabolic disease (Table 2). Subjects with BMI >25 kg/m^2^ showed a stronger reduction in BMI than those with BMI < 25 (p = 0.006). Systolic and diastolic blood pressure were reduced by 0.94 ± 8.7 mmHg and 0.31 ± 6.37 mmHg in healthy subjects, respectively, but by 9.32 ± 11.8 mmHg and −7.31 ± 7.34 mmHg in at-risk individuals. This reached statistical significance for diastolic (p = 0.009) but not systolic (p= 0.08) blood pressure. Cholesterol and LDL levels also significantly decreased in participants with elevated baseline values: cholesterol >200 mg/dl dropped by 19.7 mg/dl (p = 0.02), and LDL >130 mg/dL decreased by 16.8 mg/dl (p = 0.02).

**Table 2.**
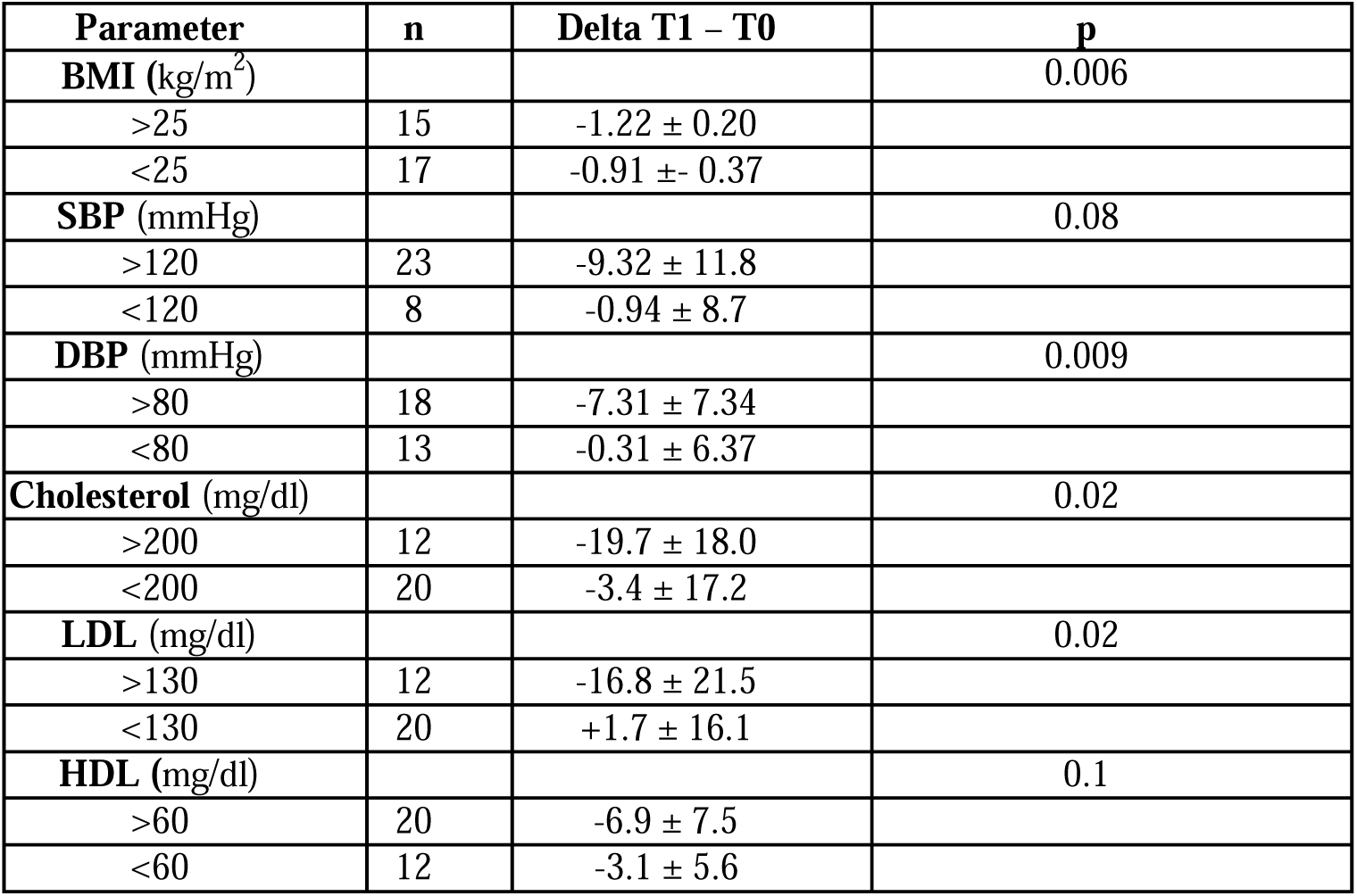
Posthoc analysis of changes in the levels of risk factors for individuals at risks of chronic disease. Not enough individuals had poor glycemic control (serum glucose level over 100 mg/dL or HbA1c levels over 5.7%). The p-values are from an independent t-test.

| Parameter | n | Delta T1 – T0 | p |
| --- | --- | --- | --- |
| **BMI (kg/m<sup>2</sup>)** |  |  | 0.006 |
| >25 | 15 | -1.22 ± 0.20 |  |
| <25 | 17 | -0.91 ± 0.37 |  |
| **SBP (mmHg)** |  |  | 0.08 |
| >120 | 23 | -9.32 ± 11.8 |  |
| <120 | 8 | -0.94 ± 8.7 |  |
| **DBP (mmHg)** |  |  | 0.009 |
| >80 | 18 | -7.31 ± 7.34 |  |
| <80 | 13 | -0.31 ± 6.37 |  |
| **Cholesterol (mg/dl)** |  |  | 0.02 |
| >200 | 12 | -19.7 ± 18.0 |  |
| <200 | 20 | -3.4 ± 17.2 |  |
| **LDL (mg/dl)** |  |  | 0.02 |
| >130 | 12 | -16.8 ± 21.5 |  |
| <130 | 20 | +1.7 ± 16.1 |  |
| **HDL (mg/dl)** |  |  | 0.1 |
| >60 | 20 | -6.9 ± 7.5 |  |
| <60 | 12 | -3.1 ± 5.6 |  |

We investigated whether the amount of 600 kcal in this program would impact individuals with different energy requirements. First, we calculated the resting energy expenditure (REE) of every participant using the validated Oxford equation ^19^. After that, we divided the 32 participants included in the MFPgroup into tertiles and compared individuals with high REE (highest tertile) vs individuals with low REE (lowest tertile). Individuals with high REE lost more weight than those with low REE (−3.9 kg vs −1.8 kg, respectively, p = 5.5×10□□). However, there were no differences in the metabolic changes in systolic or diastolic blood pressure, uric acid levels, or in parameters of coagulation, glucose, or lipid metabolism, such as total cholesterol, LDL, HDL cholesterol, or GGT (all p > 0.05). This suggests that while individuals with higher REE lose more weight due to a greater negative calorie balance, the 600 kcal per day provided in this program is able to produce similar metabolic benefits in people with lower REE. This implies that the cardiometabolic benefits of fasting in regulating glucose and lipid metabolism may be independent of the fasting induced weight loss.

### Well-being, symptoms and lifestyle

Participants answered daily questionnaires to assess their well-being, possible side effects, and lifestyle factors throughout the study. The MFP increased emotional and physical well-being, as well as quality of life evaluated using the WHO-5 index (Table 3). Energy level was higher in the MFP group after food reintroduction in comparison to baseline, but they returned to baseline after a month. Participants in the MFP group reported significantly higher levels of anxiety, cravings, headache, insomnia, and muscle weakness at baseline in comparison to the control group. Although it is unclear whether these differences existed prior to randomization, it is likely that the elevated anxiety symptoms were linked to concerns about undergoing the fasting experience. These symptoms rapidly normalised during the early days of the MFP and were no different from those experienced by the control group during the MFP and the subsequent food reintroduction. Hunger or sleep quality was not different between the two groups. Note that these findings are based on self-reported measures, which may limit their robustness.

**Table 3.** Well-being, symptoms and physical activity – questionnaires. The p-value indicates if the changes in time for a given parameter are different between the MFP group and the control (CON) group. The p-value indicates the statistical significance in a linear-mixed model where the effects of the MFP are compared to the control group at a given timepoint (*, p < 0.05 ; **, p < 0.01 ; ***, p < 0.001). Some questions were asked daily (& symbol) while some were only asked at a given timepoint. All scales are interpreted from 0 (minimum intensity) to 10 (maximum intensity) Data as mean ± SD for given timepoints.

|  | CON (T0) | CON (T1) | CON (T2) | CON (T3) | MFP (T0) | MFP (T1) | MFP (T2) | MFP (T3) |
| --- | --- | --- | --- | --- | --- | --- | --- | --- |
| EWB (0-10) & | 7.47 ± 1.88 | 7.53 ± 2.06 | 7.30 ± 2.12 | 7.63 ± 1.43 | 6.84 ± 2.50 | 8.06 ± 1.53* | 8.28 ± 1.58 | 7.63 ± 1.73 |
| WHO (0-100) | 27.5 ± 9.4 | 24.8 ± 8.7 | 23.4 ± 9.3 | 24.7 ± 8.2 | 25.9 ± 9.1 | 22.4 ± 10.2 | 18.5 ± 5.5 | 23.6 ± 10.3 |
| Physical activity (0-10) | 6.19 ± 2.49 | 5.72 ± 2.11 | 5.73 ± 2.45 | 5.76 ± 2.37 | 6.03 ± 2.09 | 5.77 ± 2.54 | 5.90 ± 2.73 | 5.37 ± 2.72 |
| Physical activity (hours/day) & | 3.64 ± 3.46 | 3.30 ± 2.94 | 3.25 ± 3.18 | 3.10 ± 2.82 | 4.39 ± 3.88 | 3.45 ± 3.04 | 4.75 ± 4.25 | 3.43 ± 3.32 |
| PWB (0-10) & | 7.31 ± 1.84 | 7.47 ± 1.87 | 7.37 ± 1.99 | 7.53 ± 1.63 | 6.78 ± 2.43 | 7.77 ± 1.78** | 8.28 ± 1.71 | 7.40 ± 2.01 |
| IPAQ (MET-min/week) | 3961 ± 3844 | 5916 ± 8170 | 2985 ± 4002 | 4491 ± 7961 | 2886 ± 2165 | 20.41 ± 21.66 | 2602 ± 3384 | 5323 ± 7932 |
| Backache (0-10) & | 1.12 ± 1.77 | 0.88 ± 1.56 | 0.60 ± 1.54 | 1.03 ± 1.76 | 2.09 ± 2.80 | 1.81 ± 2.70 | 1.03 ± 1.68 | 1.43 ± 2.21 |
| Energy level (0-10) & | 6.84 ± 2.03 | 7.25 ± 1.80 | 7.03 ± 1.96 | 7.10 ± 1.65 | 6.41 ± 2.37 | 7.10 ± 2.23 | 8.28 ± 1.71** | 7.70 ± 1.88 |
| Fatigue (0-10) & | 4.09 ± 2.96 | 3.44 ± 2.08 | 2.73 ± 2.70 | 3.45 ± 2.11 | 4.69 ± 3.04 | 3.29 ± 2.81* | 1.52 ± 2.25 | 1.97 ± 2.14** |
| Fear (0-10) & | 0.44 ± 0.88 | 0.06 ± 0.25 | 0.23 ± 0.77 | 0.17 ± 0.54 | 1.06 ± 1.83 | 0.35 ± 0.80 | 0.07 ± 0.37** | 0.30 ± 0.84 |
| Flatulence (0-10) & | 2.56 ± 2.71 | 1.69 ± 2.18 | 1.40 ± 2.04 | 1.87 ± 1.98 | 2.56 ± 2.96 | 1.55 ± 2.68 | 1.48 ± 2.29 | 1.70 ± 2.31 |
| Headache (0-10) & | 1.16 ± 2.62 | 0.50 ± 1.16 | 0.33 ± 1.03 | 0.62 ± 1.12 | 2.56 ± 3.16 | 0.23 ± 0.62*** | 0.34 ± 1.34 | 0.30 ± 1.06* |
| Hunger (0-10) & | 3.06 ± 2.94 | 1.69 ± 1.79 | 1.63 ± 2.17 | 2.03 ± 2.06 | 3.97 ± 2.92 | 3.06 ± 2.76 | 2.38 ± 2.77 | 2.83 ± 2.87 |
| Indigestions (0-10) & | 1.31 ± 2.51 | 1.06 ± 2.26 | 0.37 ± 0.72 | 0.83 ± 1.61 | 1.56 ± 2.60 | 1.16 ± 2.28 | 0.97 ± 2.24 | 0.70 ± 2.04 |
| Insomnia (0-10) & | 1.50 ± 1.88 | 1.27 ± 1.81 | 1.41 ± 1.74 | 2.41 ± 2.28 | 2.62 ± 2.34* | 2.34 ± 2.47 | 1.23 ± 2.01 | 2.07 ± 2.13* |
| Myasthenia (0-10) & | 1.16 ± 1.80 | 0.75 ± 1.68 | 0.53 ± 1.36 | 0.45 ± 0.74 | 2.31 ± 3.17 | 1.94 ± 2.24 | 0.83 ± 1.56 | 0.93 ± 1.53 |
| Ravenous appetite (0-10) & | 2.12 ± 2.66 | 0.94 ± 1.76 | 0.63 ± 1.33 | 1.17 ± 2.02 | 3.59 ± 3.05* | 2.29 ± 2.83 | 2.07 ± 2.52 | 2.53 ± 2.66 |
| Sensation of cold (0-10) & | 0.28 ± 0.68 | 0.44 ± 1.83 | 0.40 ± 1.83 | 0.80 ± 1.56 | 1.94 ± 2.97** | 0.06 ± 0.25** | 0.41 ± 1.21 | 0.30 ± 0.92** |
| Sleep quality (0-10) & | 3.03 ± 2.57 | 1.85 ± 1.99 | 2.10 ± 2.40 | 3.52 ± 2.61 | 3.53 ± 2.33 | 3.09 ± 2.83 | 2.07 ± 2.79 | 3.07 ± 2.96 |
| Backache (0-10) | 2.03 ± 1.84 | 0.88 ± 1.66 | 0.83 ± 1.7 | 1.31 ± 1.67 | 3.78 ± 2.89** | 1.77 ± 2.31 | 1.6 ± 2.25 | 2.1 ± 2.35* |
| Heartburn (0-10) | 0.69 ± 1.35 | 0.19 ± 0.64 | 0.23 ± 0.5 | 0.28 ± 0.75 | 1.44 ± 2.23 | 0.58 ± 1.48 | 0.3 ± 0.75 | 0.37 ± 0.76 |
| Joint pain (0-10) | 1.56 ± 2.18 | 1.16 ± 2.29 | 0.73 ± 1.68 | 0.79 ± 1.76 | 2.34 ± 2.68 | 1.32 ± 2.37 | 1.03 ± 1.63 | 1.6 ± 2.21 |
| Muscle (0-10) | 1.5 ± 1.65 | 0.75 ± 1.48 | 0.67 ± 1.58 | 0.72 ± 1.28 | 2.0 ± 2.29 | 2.0 ± 2.52 | 1.17 ± 1.76 | 1.13 ± 1.68 |
| Stomach ache (0-10) | 1.91 ± 2.54 | 0.62 ± 1.48 | 0.4 ± 0.81 | 0.86 ± 2.08 | 1.69 ± 2.04 | 0.81 ± 1.47 | 1.0 ± 1.88 | 1.0 ± 1.74 |
| Stress (0-10) | 4.44 ± 1.93 | 3.59 ± 2.15 | 3.93 ± 2.16 | 3.79 ± 2.29 | 5.5 ± 2.16* | 4.03 ± 2.51 | 3.97 ± 2.51 | 4.2 ± 2.23 |
| PSQI (0-20) | 7.59 ± 2.75 | 6.66 ± 2.91 | 5.73 ± 2.56 | 6.16 ± 2.92 | 8.50 ± 3.31 | 7.68 ± 3.02 | 6.33 ± 2.54 | 6.71 ± 2.67 |

### Serum metabolomics reflects remodelling of energy metabolism

NMR metabolomics revealed that the MFP caused marked metabolic changes, which persisted during the food reintroduction and returned to baseline after a month (Figure 2). We observed that concentrations of 24 metabolites were significantly decreased in the MFP in comparison to the control group, and that 9 metabolites were increased. In line with expectations from metabolic switching, strong increases were observed for the ketone bodies acetoacetic acid and 3-hydroxybutyric acid after the MFP. Strongly decreased metabolites include amino acids (i.e., proline, alanine, glutamine, glycine, methionine, lysine, serine, threonine, histidine). Other notably decreased metabolites include glycoprotein acetyls A, a known biomarker of chronic inflammation ^20^. Altogether, this serum metabolomics analysis reflected a strong metabolic remodelling caused by the fasting intervention, but also a return to baseline after a month.

**Figure 2.**
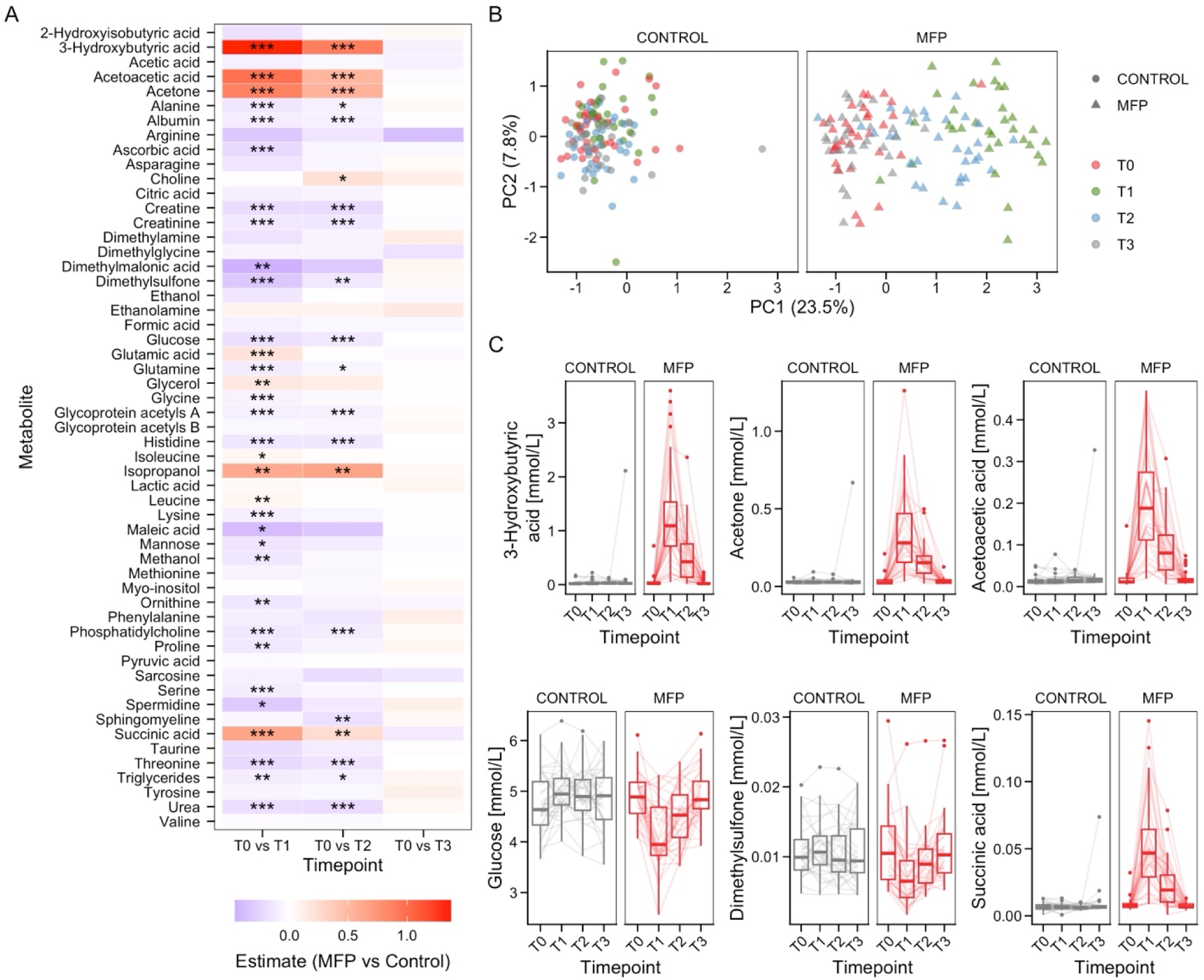
Serum metabolomics analysis of the effects of the MFP on serum metabolite composition. **A:** Heatmap showing the changes in the levels of metabolites between the MFP group and the control group at different timepoints. The colour scale shows the estimate of the difference between the MFP group and the control group in a linear mixed model with individuals as random effects. The p-value resulting from these models was adjusted for multiple comparisons using the Benjamini-Hochberg procedure (*, p < 0.05 ; **, p < 0.01 ; ***, p < 0.001). **B:** Principal Component Analysis (PCA) shows the structure of the variance between these groups and timepoints. The PCA was done for all samples together, but the grouping of samples are visualised facetted for the sake of clarity. **C:** Time trajectory for the abundance of 6 metabolites with the lowest p-values.

### Shotgun metagenomics reveals foraging of host-derived substances

Changes in the gut microbiota were investigated by performing shotgun metagenomics on participants’ faecal samples. Both alpha diversity analysed with the Shannon index (Figure 3A) and beta diversity analysed with Bray-Curtis dissimilarity remained unchanged (Figure 3B). Limited changes were observed with only 11 species having their relative abundance statistically significantly changed at T1 of MFP versus T1 of controls (as assessed by mOTUs, Figure 3C). No changes were observed at T3 compared to T0, indicating that the effects on the gut microbiota were transitory.

**Figure 3.**
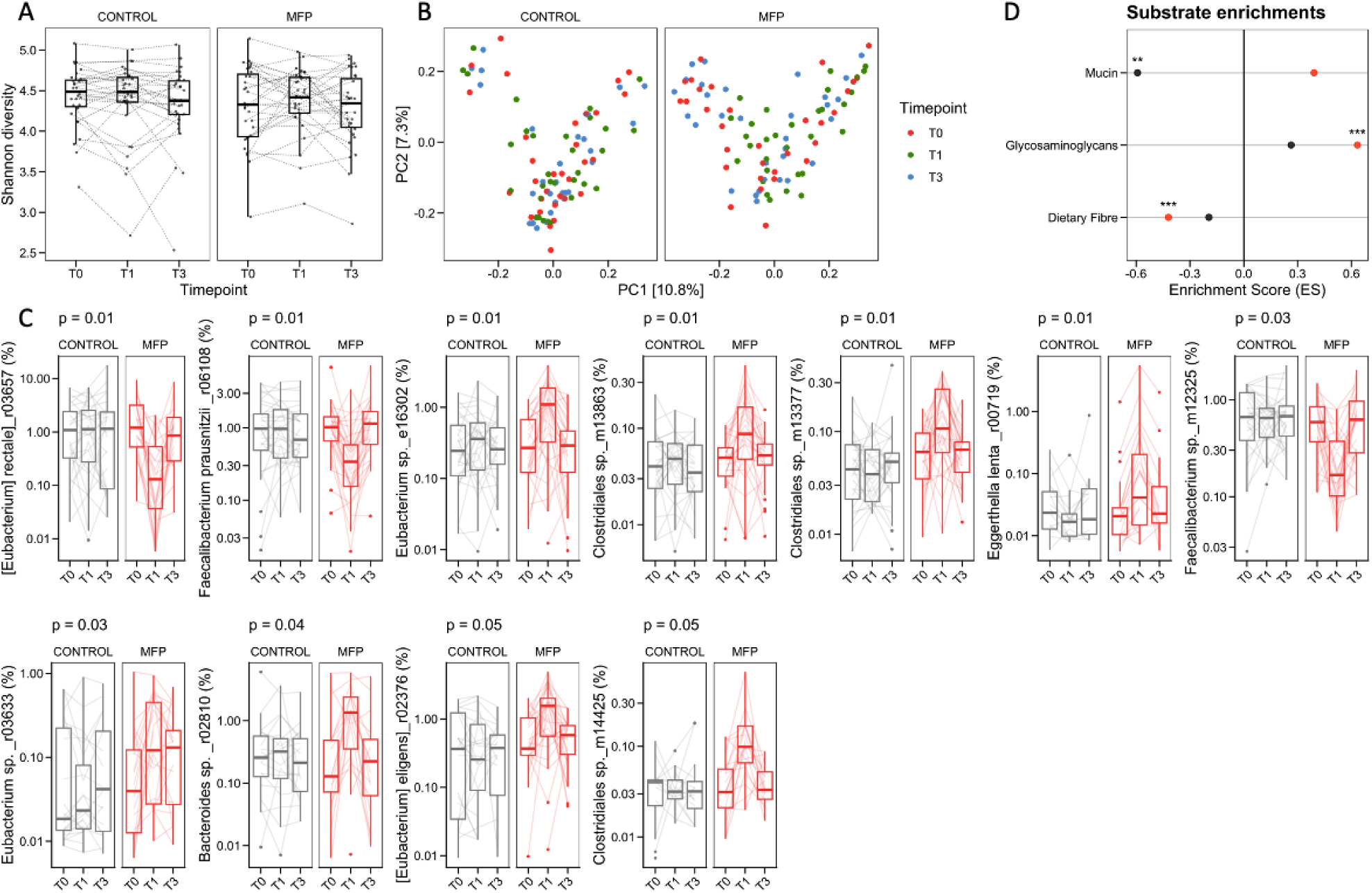
Changes in faecal microbiota composition and function. **A:** Alpha diversity was computed as Shannon diversity. **B:** The Principal Coordinate Analysis (PCoA) of microbial communities is based on Bray-Curtis dissimilarities, coloured by timepoints and faceted by study arms. **C:** Individual changes in abundance for the 11 bacteria species with the p-values <0.05 in the evaluation of the effect of the MFP (T1 vs T0). **D.** All CAZyme families were additionally subjected to gene-set enrichment analysis (GSEA) to evaluate substrate enrichments (p-values are indicated as start symbols, * p<0.05 ; ** p<0.01 ; *** p<0.001). Generalized fold changes were calculated using linear mixed models (LMMs) with random intercepts for the individuals.

Changes in energy metabolism in the gut microbiota were even clearer. We analyzed substrate preference derived from CAZyme abundances quantified from metagenomes ^21^. A total of 52 CAZymes had their abundance significantly changed at T1 of MFP versus T1 of controls, but none were persistently changed at T3. This reflected a transient reduction in the abundance of CAZymes that metabolise dietary fibre concomitant with an increase in CAZymes involved in host-derived glycosaminoglycan foraging. Gene-set enrichment analysis (GSEA) to evaluate substrate enrichments suggested that the gut microbiome switched metabolism and started foraging host-derived substances during the MFP intervention (Figure 3H).

### Comparison with 5 days of prolonged fasting

We compared the effects of the MFP to the effects of 5 days of PF according to the Buchinger Wilhelmi protocol (75 - 250 kcal/day). In order to allow for fair comparisons between groups, we matched each of the 32 participants from the MFP group to individuals with similar sex, age, blood pressure, triglycerides, BMI, waist circumference from a total of 400 patients who performed 5 days of fasting in the Buchinger Wilhelmi clinic in 2016 ^8^. Effect sizes of the MFP or 5 days of fasting were comparable for most clinical parameters (Figure 4A and Table S6). Effects of the MFP were slightly more pronounced than 5 days of prolonged fasting for the changes in the anticoagulative effects and the glucose levels. The latter is related to the glucose content of the supplementation, higher in PF and then MFP. Altogether, this shows that the MFP has comparable beneficial metabolic effects to PF.

**Figure 4.**
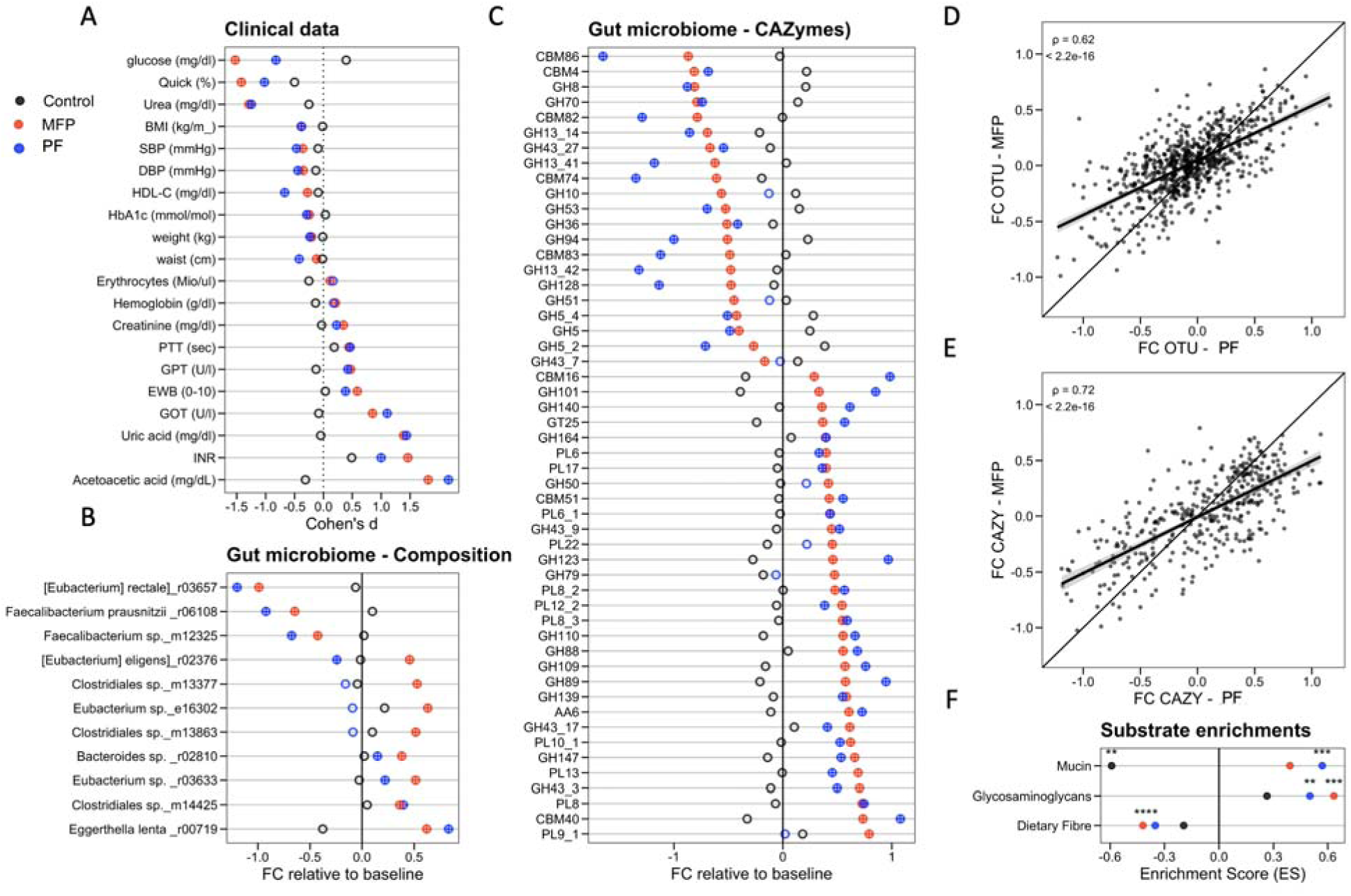
Metabolic changes caused by the MFP were comparable to those of a 5-day prolonged fasting. **A.** Comparison of the effect size estimated from the effects of 5-day MFP in comparison to the control group at T1 for parameters showing statistically significant changes in their levels at T1. These effects are also compared to 5 days of prolonged fasting in 32 individuals matched based on their age, sex, body weight and triglyceride levels out of 400 participants who fasted according to the Buchinger Wilhelmi fasting programme (250 kcal/day) in 2016. **B:** Dot plot shows fold changes in differential abundance in microbiota composition before and after the MFP intervention in comparison to PF in another study. **C:** Differential CAZyme family abundance in comparison to PF in another study. **D:** Correlations between fold changes in relative abundance for microbiota composition. **E:** Correlations between fold changes for CAZyme family abundance. **F**. All CAZyme families were additionally subjected to gene-set enrichment analysis (GSEA) to evaluate substrate enrichments (p-values are indicated as start symbols, * p<0.05 ; ** p<0.01 ; *** p<0.001). Parameters are ordered by MFP effect sizes. Statistical significance is indicated with crossed circles (p<0.05).

We compared the directionality of the changes in gut microbiota composition with the changes caused by PF we observed in another study ^22^. The changes induced during PF are comparable to those induced by the MFP (Figure 4B), as shown by the statistically significant correlation between generalized fold changes for each mOTU measured in both interventions (Figure 4D). The changes in CAZy families were highly correlated (Figure 4E) and thus comparable to those observed during PF (Figure 4B). Gene-set enrichment analysis (GSEA) suggested that the gut microbiome switched metabolism in a comparable way to what is observed during PF (Figure 4F).

## DISCUSSION

Large research efforts are currently undertaken to find pharmacological and non-pharmacological interventions for maintaining body weight and blood pressure within recommended limits, in order to reduce the high incidence of chronic metabolic diseases such as type 2 diabetes or coronary heart disease ^23^. The possibility to modulate cardiovascular risk factors triggers the need for proactive healthcare policy planning ^24^. The MFP evaluated in the present RCT demonstrates that short fasting periods performed at home are an effective strategy for reducing body weight and improving the cardiometabolic profile.

Compared to pharmacological approaches for weight loss and metabolic normalisation, such as GLP-1 agonists ^25^, fasting strategies generally present with only mild side effects, particularly when conducted under professional supervision, either in specialised institutions or for shorter periods through online platforms, as is the case of MFP ^8^. Unlike GLP-1 agonists, which are associated with significant costs ^26^, non-pharmacological strategies like MFP empowers individuals to explore and manage on their own dietary strategies enhancing a sense of self-efficacy and safety ^27^. The alternance of MFP with a normal diet triggers well-being and diminishes cravings often associated with continuous caloric restriction. MFP offers health benefits and is compatible with daily life ^28^. Short and periodically repeated interventions also prevents from developing with its deleterious effects on the immune system and wound healing ^29^. Nevertheless, the risk of falling back into old lifestyle habits that led to poor metabolic health cannot be fully eliminated.

The mechanisms through which the MFP intervention may confer health benefits seem to be multifaceted and deeply interconnected. The loss of body weight by itself could explain a decrease in chronic inflammation ^30^. The clinical and metabolic changes caused by the MFP were comparable to those observed after PF (i.e. total of 5 days of fasting at 250 kcal/day). The changes caused by the metabolic shift into ketosis fasting are known to contribute to beneficial changes in nutrient-sensing signalling pathways ^31,32^, which play crucial roles in regulating cellular processes related to metabolism, growth, autophagy, stem cell-based regeneration and ultimately longevity ^14^.Therefore, by modulating these pathways, our MFP holds substantial potential for enhancing public health.

A single cycle of the MFP showed maintenance of weight loss after a month in predominantly normal weighted individuals, while other parameters returned to baseline. The easy implementation makes it a promising candidate for multiple-cycle programs, potentially mirroring the enhanced outcomes seen with Fasting Mimicking Diets (FMD). In humans, three cycles of a monthly 5-day FMD in a RCT involving 100 healthy subjects resulted in weight loss of on average 2.6 ± 2.5 kg, decreased BMI, total body fat, trunk fat and reduced waist circumference ^33^. It also reduced the need for medication and improved glycaemic control in primary care for type 2 diabetes ^34^. A study involving 24 healthy volunteers confirmed the glucose-lowering effect of a 5-day FMD and also demonstrated reductions in IGF-1 alongside improved insulin sensitivity ^35^. A recent study even showed that three cycles of FMD had the potential of slowing down markers of biological ageing ^36^. Longer follow-up including different frequencies of MFP use will be needed to understand whether this MFP can be used to treat chronic diseases.

Our study shows that the MFP is an effective method to address risk factors for cardiometabolic diseases. A main strength of our study is the joint collection of patient blood markers, metagenomic and metabolomic data in a longitudinal design surveyed under randomised controlled clinical conditions. This helped us demonstrate that this MFP can exert beneficial health effects by eliciting mechanisms which are known hallmarks of the fasting metabolism. We even noted that participants in the MFP group with higher baseline values above the norm ranges experienced more significant improvements in metabolic parameters compared to those whose values were within the normal range. This was suggested in another study where metabolic markers like fasting glucose, triglycerides, CRP, and cholesterol levels, which were within the normal range at baseline, did not show significant changes after three cycles of FMD in a randomised comparison ^33^. The same is true for the PF ^8^, showing that at-risk individuals may benefit more from the intervention. Another strength of our program is that participants received educational material to improve their lifestyle including instructions to do more physical activity. It can also be seen as a limitation, because even if the control group received similar advice and could be used to differentiate the effects of the MFP from the effects of the accompaniment, it is not always practical to disentangle the contribution of the numerous factors which compose such a lifestyle intervention. Other limitations include the focus on individuals who were mostly metabolically healthy, making it challenging to predict the potential benefits or difficulties that might arise in patients with underlying conditions. Therefore, further research is needed to explore the effects of MFP in patients with metabolic conditions such as hypertension or type 2 diabetes, or fatty liver disease chronic diseases.

This new programme is in line with the need for lifestyle interventions which substantially decrease healthcare costs ^37,38^. At-home interventions are accessible to a broad population, and are compatible with individuals’ daily lives, thereby increasing the likelihood for long-term adherence. This could be the case when individuals are confined at home in isolation such as during the Coronavirus-19 disease (COVID-19) pandemic ^39,40^ or other reasons.

In conclusion, the results of this RCT show that the MFP is an effective intervention for reducing body weight, lowering blood pressure and inducing a metabolic switch to ketosis thus inducing similar beneficial effects than those observed in other fasting interventions. The improvements in cardiovascular and metabolic risk factors, especially in subjects with elevated baseline levels of these markers, are of significant value for non-pharmacological public health strategies.

Future investigations should prioritise the study of at-risk patient groups to understand the efficacy of MFP in reducing lifestyle-related diseases, assess long-term effects, and explore the fasting-like effects to gain insight into its broader preventive and therapeutic benefits for promoting health and longevity.

## MATERIALS AND METHODS

### Subjects

The Fastreset study was designed as a RCT with two parallel study arms. Subjects were randomised either to five days of MFP followed by four days of food reintroduction or to their usual diet (Figure 1B). The study protocol was approved by the Ethics Committee of the Medical Association of Baden-Württemberg, Stuttgart, Germany (approval number: F-2023-014). The study was registered at ClinicalTrials.gov (NCT05821660) prior to the start of patient recruitment. The study was conducted in accordance with the principles of the Declaration of Helsinki and the standards of Good Clinical Practice. The study visits were carried out on an outpatient ambulatory basis between June and August 2023 at the Buchinger Wilhelmi Clinic in Überlingen (Baden-Württemberg, Germany). Written informed consent was obtained from all participants.

### Dietary intervention

The MFP used was the Buchinger Wilhelmi FASTING BOX (Buchinger Wilhelmi Development und Holding GmbH, Germany) which includes ready-made meal replacements to be consumed over a five-day period. The ingredients are organic, plant-based products. The 24 items include vegetable soups as the main supplement, high-quality oils (linseed and algae oil) rich in omega 3 polyunsaturated fatty acids, chickpea puree, apple puree, honey and cashew nuts. The MFP provides on average 623 kcal per day, consisting of 9.8 grams (6.2 % of caloric intake) of proteins, 41.5 grams (26.6 %) of carbohydrates, and 44.7 grams (64.7 %) of fat (Table S1). The aim of the MFP regimen is to achieve a steady reduction in calorie intake while limiting carbohydrate consumption to less than 50 g and proteins to less than ∼10 g per day. The MFP includes a food supplement containing Tricalcium citrate, potassium citrate, magnesium citrate, zinc citrate trihydrate, sodium chloride. Furthermore, herbal teas, soup essences and herbal extracts are included as refreshment options, along with the recommendation to drink at least 2.5 L non-caloric, caffeine-free beverages daily. The consumption of water and herbal tea is not restricted. A detailed meal plan describes the recommended daily food intake and reflects the energy, fat, carbohydrate and protein content (Table S1).

The 5-day program begins with a transition day. This is the only day when a morning serving is provided. On the following four MFP days, subjects skip the morning meal inducing a time-restricted eating pattern. The entire MFP programme is gluten-free and contains only a very restricted diversity of food items. The MFP was followed by four days of gradual food reintroduction from 800 to 1600 kcal per day. Half of the intervention group received a standard food reintroduction (n=16) and half received a ketogenic food reintroduction (n=16) (Table S2). The food was freshly cooked, organic, plant-based meals to take home after their daily measurements. Subjects in the control group were advised to stick to their usual diet.

### Participants

Participants were recruited from the local area of Überlingen, Germany. Information about the study was offered in social media and per mail. An online questionnaire was used to recruit volunteers. If they appeared eligible, potential candidates were invited to an online presentation of the study. They were then invited to an assessment with the study physician, who checked the inclusion and exclusion criteria and asked for written informed consent. Women and men between 18 and 80 years were eligible. The following criteria led to exclusion: (1) intake of antibiotics up to 2 months prior the study; (2) medicated high blood pressure; (3) diagnosed hyperuricemia; (4) diagnosed diabetes mellitus type I and II; (5) diagnosed kidney stone; (6) active malignant diseases; (7) known substance addiction; (8) pregnancy or breastfeeding; (9) diagnosed with cachexia, anorexia, nervosa, advanced kidney, liver or cerebrovascular insufficiency; (10) inability to sign the informed consent and (11) participation in another study. The participants received no fee for their study participation.

### Measurements

After a thorough physical examination by the study physician participants were randomised to either the intervention or control group. In order to facilitate the conduction of the study by the clinical centre, the study was conducted in two phases, each comprising 32 participants from both the control and intervention groups, with a one-month interval between them. For eleven consecutive days, the participants were examined daily in the morning. Four main visits were conducted: T0 at baseline (BL) before the start of the intervention, T1 in the morning of the 7th day after the five-day MFP (end MFP), T2 in the morning of the 11th day after the subsequent four-days of food reintroduction (FR) and T3 at follow-up one month later.

### Endpoints

The changes in blood pressure and body weights are considered as primary endpoints. Changes in lipid and glucose metabolism, inflammatory parameters, total antioxidant capacity, and well-being were also analysed using linear mixed models with adjusted p-values < 0.05 being considered significant. All other parameters are considered as exploratory endpoints.

### Clinical examination

Clinical parameters were measured daily by trained research assistants. Body weight was evaluated with participants lightly dressed, utilising a Seca 704 (Seca, Hamburg, Germany). Height was measured using the same device. Blood pressure and heart rate were measured twice on the non-dominant arm while seated, after a resting period, using a boso Carat professional (Bosch und Sohn GmbH u. Co. KG, Jungingen, Germany). Waist circumference was determined using a tape, which was placed halfway between the lowest rib and the iliac crest (openmindz GmbH, Heidelberg, Germany). Acetoacetic acid was self-measured twice per day in the first morning and last evening urine using Ketostix (Bayer, Leverkusen, Germany).

### Blood parameters

Blood samples were collected at all four main visits (baseline, end of fasting, end of food reintroduction period, follow-up) by trained medical assistants after participants had fasted overnight (Figure 1B). Lipid, glucose, kidney, liver and inflammatory parameters were determined as described in a previous study ^41^. The antioxidative capacity of water-soluble substances (ACU) and lipid-soluble substances (ACL) was determined by photochemiluminiscence in serum and lipidperoxide by photometry in EDTA plasma.TNF-alpha was measured in serum by chemiluminiscence immunoassay.

### Questionnaires

Self-reported data was documented by completing online questionnaires in the Buchinger Wilhelmi Amplius app. The participants received instructions and further information via this app to enable them to participate in this programme remotely. Adverse events were recorded in a report form by study physicians.

Participants reported the following outcomes on all four main visits (T0-T3): The Well-being Index World Health Organization 5 (WHO-5) ^42^, International Physical Activity Questionnaire (IPAQ) - short form, Pittsburgh Sleep Quality Index (PSQI) ^43^, Short healthy eating index (sHEI) ^44^, and Questionnaire of highly processed food consumption (sQ-HPF) ^45^. Furthermore, lifestyle habits such as smoking, and alcohol consumption were documented. Stress level was rated on a visual scale from 0 (none) to 10 (extreme), and energy level from 0 (weak) to 10 (full of energy). Alcohol consumption was recorded as glasses of beer (0.5 l; 2 units of alcohol), wine (0.25 l; 2 units of alcohol) or spirits (0.02 l; 1 unit of alcohol) per week and added up to the number of drinks per week.

Additionally, participants self-reported daily their emotional and physical well-being as well as energy level on visual scales from 0 (very bad) to 10 (excellent). Sleep disturbances (0, none to 10, very strong) and sleep quality (0, calm to 10, restless) were reported. Further, participants rated whether they had cravings, felt bloated or cold (0; strongly disagree to 10; strongly agree). Symptoms such as fatigue, muscle weakness, back pain, headache, hunger, anxiety and digestive disorders on visual scales from 0 (none) to 10 (maximum) were evaluated. Self-perceived physical activity was recorded on a visual scale from 0 (inactive) to 10 (very active) and the actual activity time was captured in hours per day.

### Faecal microbiota shotgun metagenomics

Three faecal samples were collected using EasySampler Stool Collection Kit (GP Medical Devices, Holstebro, Denmark) at T0, T2 and T3. Samples were stabilised by DNA/RNA ShieldTM Reagent, Zymo Research Europe GmbH, Freiburg, Germany and stored at −20°C. The samples were processed and analysed by CeGaT, Tübingen, Germany.

Genomic DNA samples were profiled with shotgun metagenomic sequencing. Sequencing libraries were prepared with the Illumina preparation kit (M) Tagmentation (Illumina, San Diego, CA) with 100 ng genomic DNA following the manufacturer’s protocol. Q30 value of sequencing was >90.25%. The final library was sequenced on the NovaSeq 6000® (Illumina, San Diego, CA) platform.

Metagenomics shotgun sequencing data processing was performed as described in Ducarmon et al. ^22^. Raw reads underwent quality filtering using bbduk (v38.93): 1) low-quality trimming (qtrim=rl, trimq=3), 2) discarding low-quality reads (maq=25), 3) adapter removal, and 4) length filtering (ml=45). Taxonomic profiling used mOTUs (v3.1), while functional profiling filtered out human genome reads via kraken2 (v2.1.2) against hg38, then mapped to a reduced GMGC human gut catalogue. Profiling of carbohydrate-active enzymes was done using Cayman ^21^.

Statistical analysis of metagenomic data was done in the statistical programming language R. Alpha-diversity was assessed using in phyloseq. Differences between baseline and other timepoints were statistically evaluated using a linear mixed model (LMM). Beta-diversity was determined using Bray-Curtis dissimilarity. Differential abundance analysis was done using LMM in SIAMCAT (v2.5.1), with participant-specific random effects. Features with <10% prevalence were excluded. Pseudocounts varied and were 1e-4 for taxonomic analysis or 0.01 RPKM for CAZy. FDR correction was applied to p-values, with significance defined as adjusted p < 0.05.

Functional analyses in gut metagenome were done as described in Ducarmon et al. ^22^. Gene-set enrichment analysis (GSEA) using fgsea identified higher-level CAZy substrates.

### Serum metabolomics

Serum metabolomics was performed by lifespin GmbH (Regensburg, Germany). Sample preparation was done from 350 μl of serum mixed with 350 μl of aqueous buffer. The buffer consists of H_2_O p.A., 0.1 g/l NaN_3_, 0.067 mol/l Na_2_HPO_4_, 0.033 mol/l NaH_2_PO_4_ (pH: 7.15 ± 0.05), 5% D_2_O field-lock substance and an internal standard (6 mM pyrazine) for quantification. From this mixture, 600 μl were transferred into a 5 mm Bruker NMR tube and sealed with barcode-labelled lids. The final NMR samples were stored after preparation at refrigerator temperature until measurement, not more than 24 h. NMR measurement was performed on a Bruker AVANCE NEO 600 MHz (measuring method 1D 1H noesygppr1d_d20, NS=16, T=310K).

The spectra obtained were Fourier transformed using TopSpin software (version 4.0, Bruker Biospin, Germany). All spectra were automatically phased and subjected to baseline correction. Subsequently, the spectra were analyzed using the proprietary lifespin Profiler software (version 1.4_Blood) and a quantitative metabolite list was generated.

### Statistical analysis

Based on previously collected data for changes in systolic blood pressure ^46^, we calculated that a sample size of 32 participants per arm is required to detect a decrease in 1 mm/hg per day with three groups and an effect size of 0.33 with 90% power and an alpha error probability of 0.05 with an ANOVA test including repeated measurements (G*Power version 3.1.9.7). Randomisation was performed with R, with a custom script splitting individuals into two groups randomly with equal mean age and weight.

Statistical significance for the primary endpoints was assessed using LMM. Extraction of 32 closest neighbors of our participants, from subjects of our study with a cohort of 1422, was done using the NearestNeighbors class from the module neighbors of sklearn (1.3.2). The distance computed was the Euclidean, based on the following normalized parameters: BMI (kg/m²), age, SBP, waist circumference, triglycerides (Table S3). All statistics and tables were generated with python using the packages statsmodels (0.14.0) and scipy.stats (1.11.3). Differences were calculated using unrounded values to maintain accuracy, with rounded values presented for clarity. For the baseline comparison we used a t-test with adjustment for equality of the variance depending on the outcome of a Levene test. All other comparisons were measured using mixed models with the participant ID considered as a random effect. P-values less than 0.05 were considered to be statistically significant.

## ADDITIONAL INFORMATION

## ACKNOWLEDGMENTS

We are grateful for the continuous support of the whole staff of Buchinger Wilhelmi Clinics.

## CODE AVAILABILITY

The code used to perform the statistical analysis is available upon request. Scripts used to process the shotgun metagenomics data are available on GitHub.

## AUTHOR CONTRIBUTIONS

FG, FWT, and RM conceptualized and designed the study. AH contributed to statistical analyses, with support from RM. SS, SH and RH performed the metabolomics analysis. QRD and GZ performed the shotgun metagenomics analysis. BMT and MRM contributed to data analyses and interpretations. RM, FG and FWT drafted the initial version manuscript. RM wrote the final version of the manuscript. All authors reviewed, edited, and approved the final version of the manuscript.

## DATA AVAILABILITY

Raw metagenomic data will be made available upon acceptance of the manuscript under accession number PRJEB75353 at ENA.

## SUPPLEMENTARY MATERIAL

**Figure S1.**
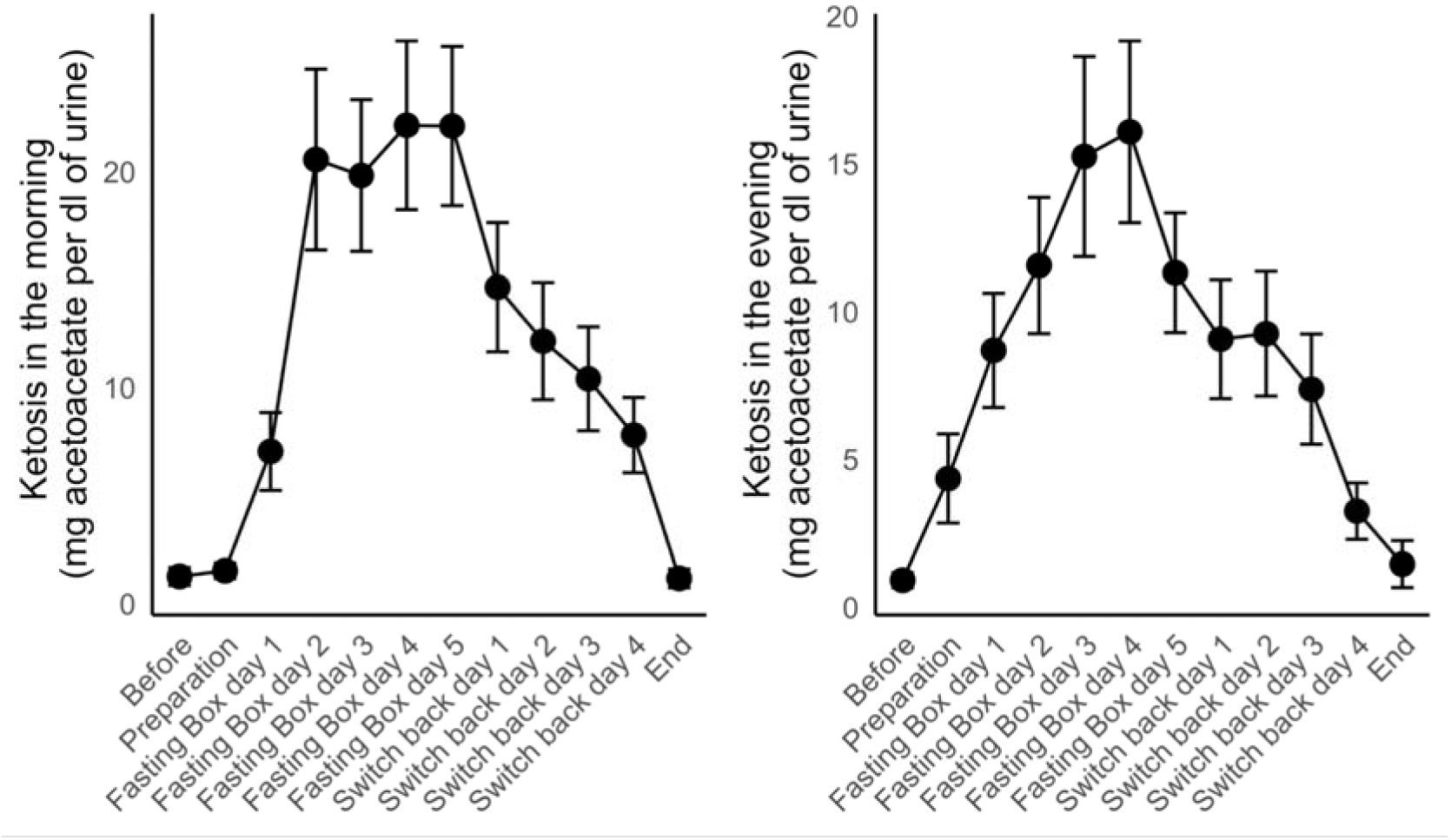
Ketonuria in the group of 32 participants in the MFP arm. Ketosis was not detectable in the control group. Data as mean ± SD.

**Table S1.** Meal plan during the 5-day MFP and the energy and macronutrient content provided.

|  | **Day 1** | **Day 2 -4** | **Day 5** |
| --- | --- | --- | --- |
|  | **Transition** | **Fast Reset Day** | **Restart** |
| **Breakfast** | Yoghurt 1.5 % fat<br>Cashews<br>Oil | - | - |
| **Lunch** | Soup,<br>Oil | Soup<br>Chickpea puree<br>Oil | Apple puree<br>Cashews<br>Oil |
| **Dinner** | Soup<br>Oil | Soup<br>Chickpea puree<br>Oil | Soup<br>Oil |
| **During the day** | Mineral capsule | Mineral capsule | Mineral capsule |
|  | Herbal tea | Herbal tea | Herbal tea |
|  | Herbal extract | Herbal extract | Herbal extract |
|  |  | Soup essence | Soup essence |
| **Energy (kcal)** | 648.4 | 600 | 620.7 |
| **Fat (g)** | 44.6 (61.9 %) | 47.0 (70.5 %) | 42.5 (61.6 %) |
| **Carbohydrate (g)** | 43.8 (27.0 %) | 32.8 (21.9 %) | 47.9 (30.9 %) |
| **Protein (g)** | 16.1 (9.9 %) | 7.0 (4.7 %) | 6.2 (4.0 %) |

**Table S2.**
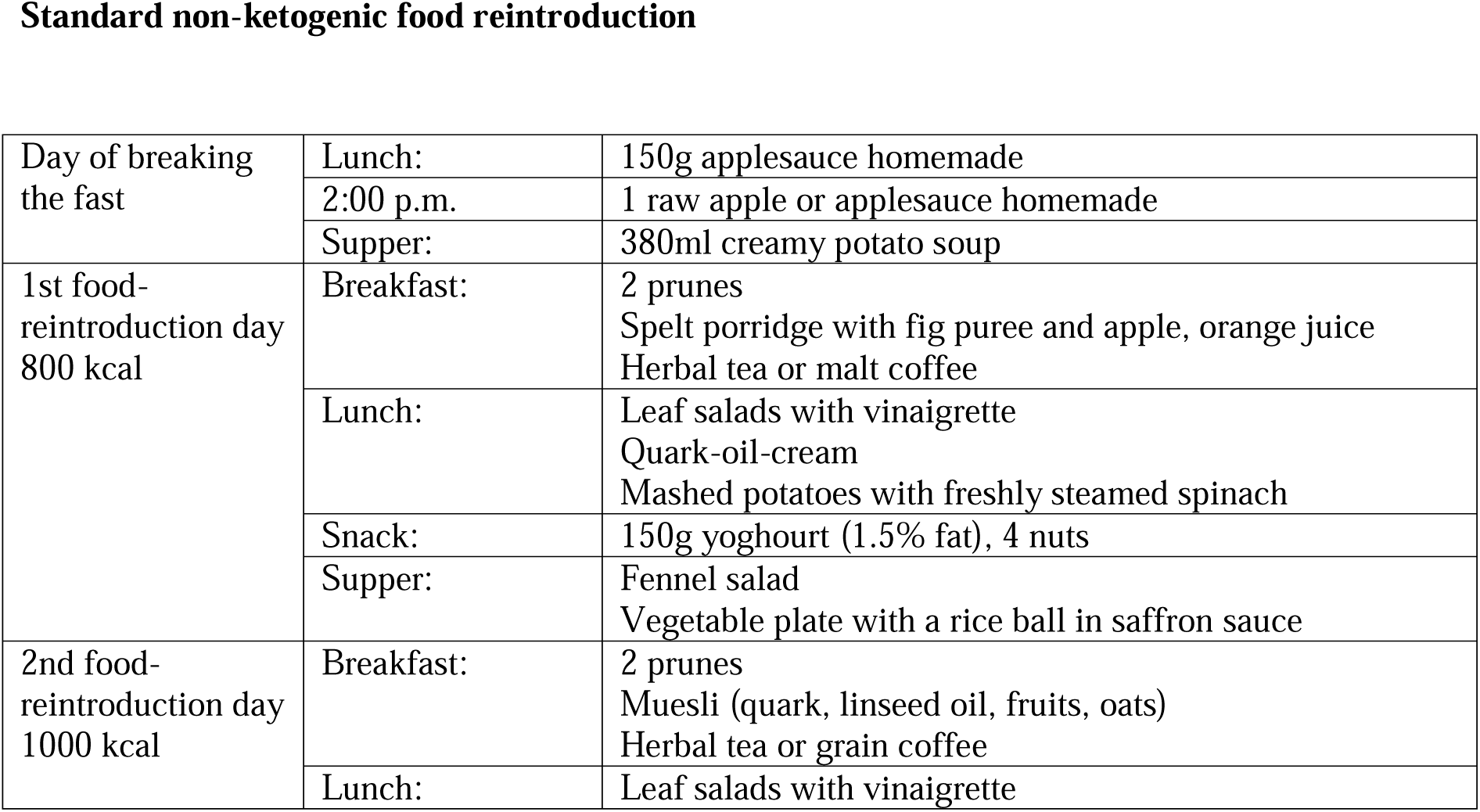
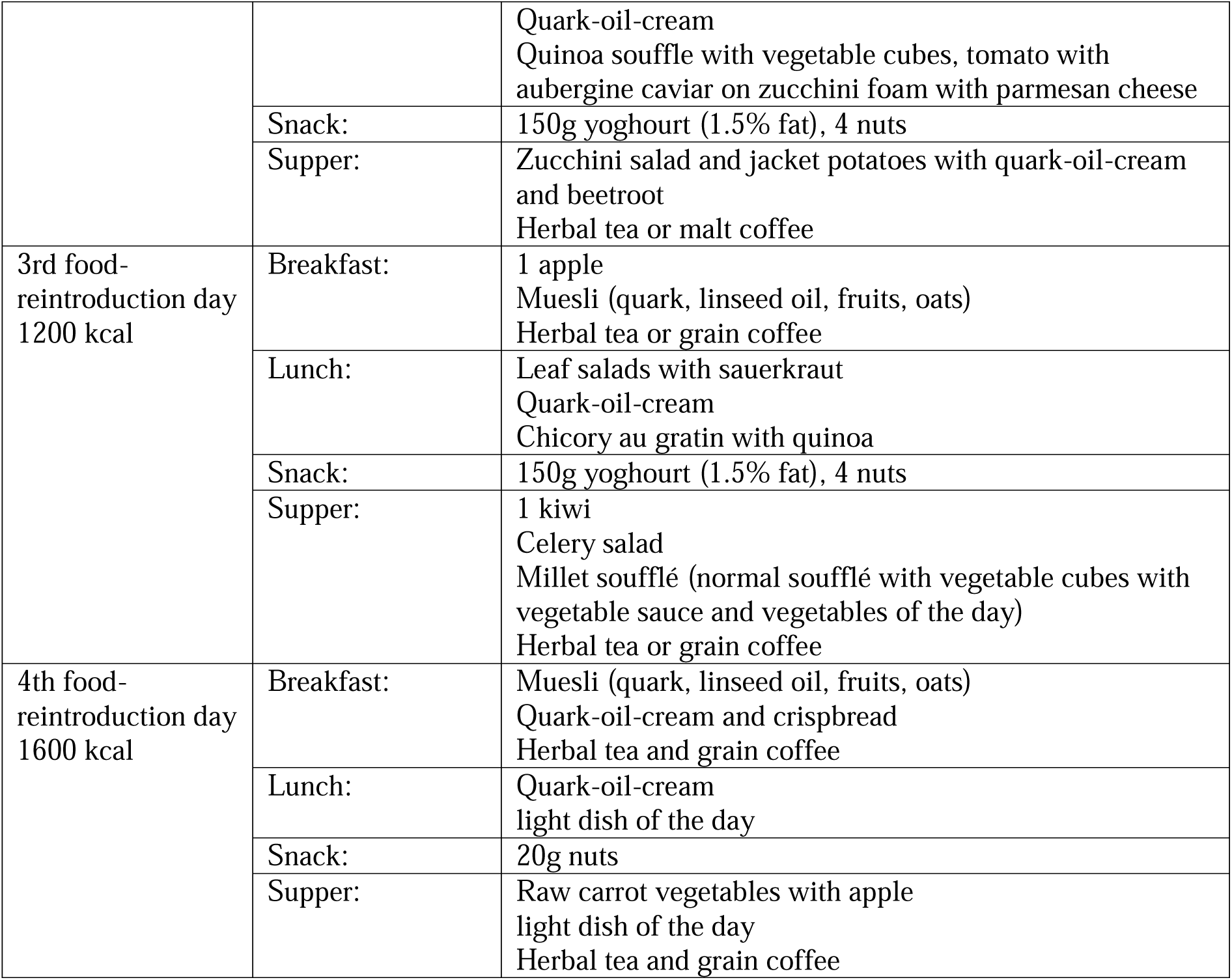
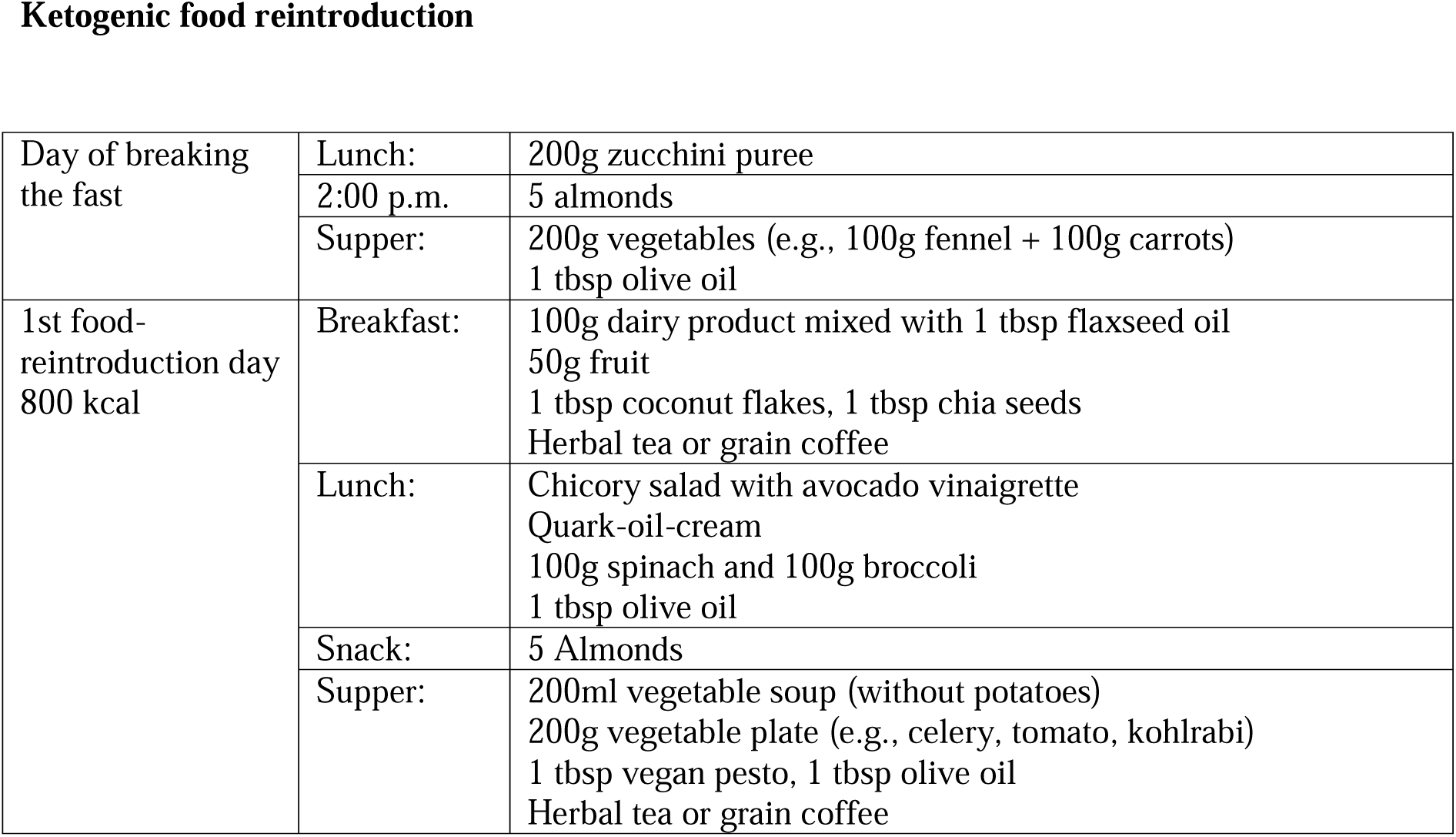
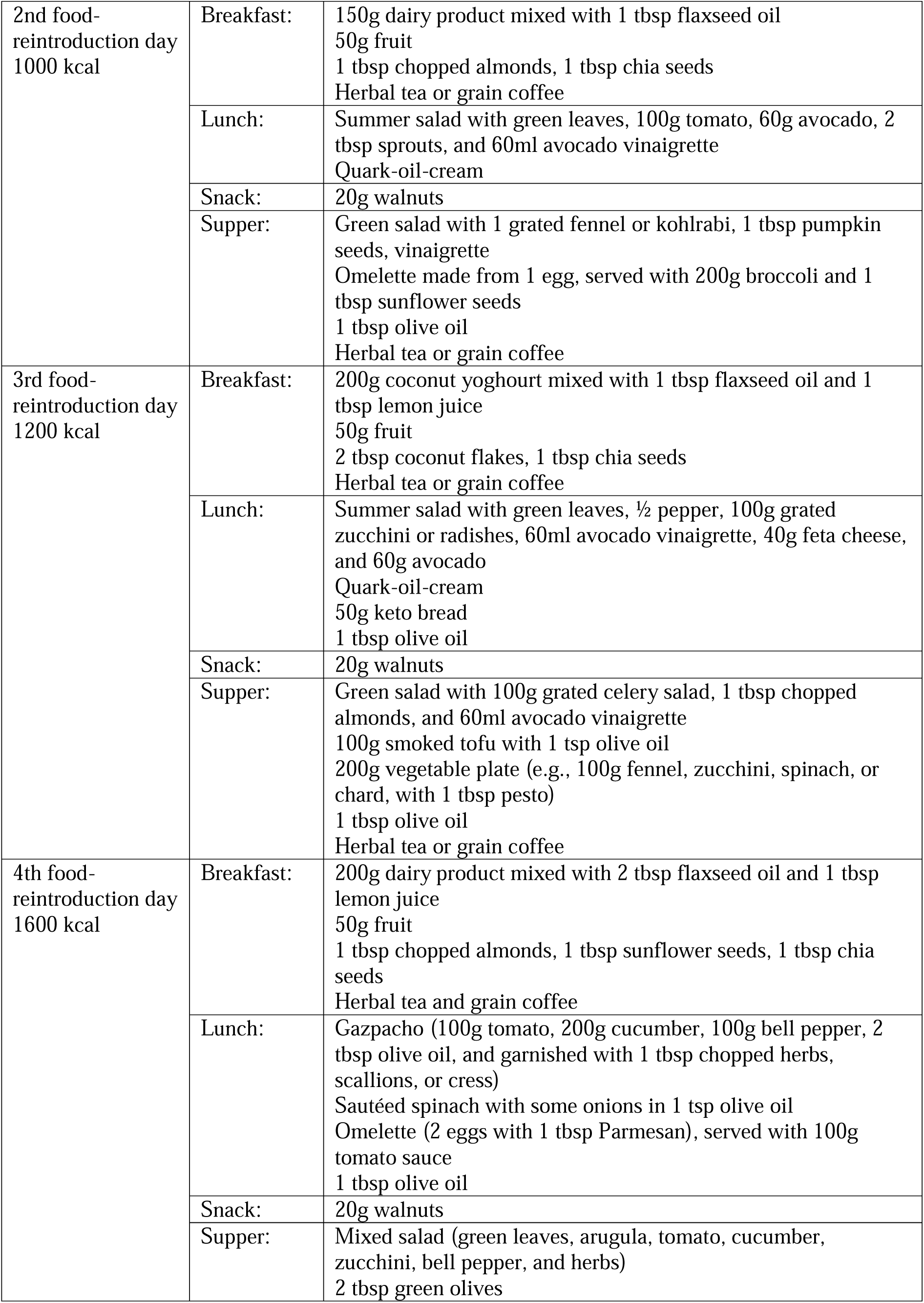
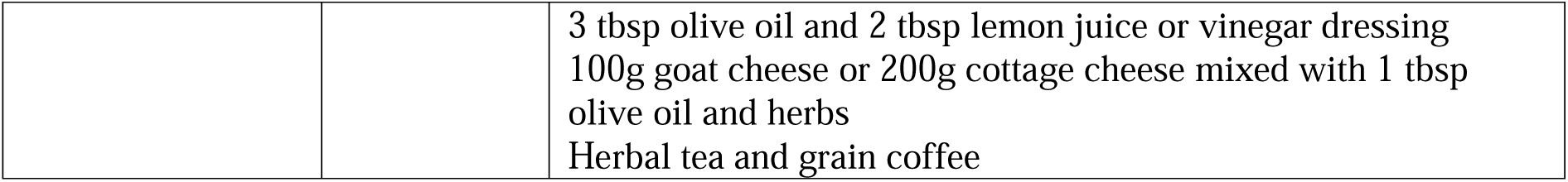
Food reintroduction for the ketogenic group and the non-ketogenic group.

**Standard non-ketogenic food reintroduction**
|  |  |  |
| --- | --- | --- |
| Day of breaking the fast | Lunch: | 150g applesauce homemade |
|  | 2:00 p.m. | 1 raw apple or applesauce homemade |
|  | Supper: | 380ml creamy potato soup |
| 1st food-reintroduction day<br>800 kcal | Breakfast: | 2 prunes<br>Spelt porridge with fig puree and apple, orange juice<br>Herbal tea or malt coffee |
|  | Lunch: | Leaf salads with vinaigrette<br>Quark-oil-cream<br>Mashed potatoes with freshly steamed spinach |
|  | Snack: | 150g yoghourt (1.5% fat), 4 nuts |
|  | Supper: | Fennel salad<br>Vegetable plate with a rice ball in saffron sauce |
| 2nd food-reintroduction day<br>1000 kcal | Breakfast: | 2 prunes<br>Muesli (quark, linseed oil, fruits, oats)<br>Herbal tea or grain coffee |
|  | Lunch: | Leaf salads with vinaigrette |
|  |  | Quark-oil-cream<br>Quinoa souffle with vegetable cubes, tomato with aubergine caviar on zucchini foam with parmesan cheese |
|  | Snack: | 150g yoghourt (1.5% fat), 4 nuts |
|  | Supper: | Zucchini salad and jacket potatoes with quark-oil-cream and beetroot<br>Herbal tea or malt coffee |
| 3rd food-reintroduction day<br>1200 kcal | Breakfast: | 1 apple<br>Muesli (quark, linseed oil, fruits, oats)<br>Herbal tea or grain coffee |
|  | Lunch: | Leaf salads with sauerkraut<br>Quark-oil-cream<br>Chicory au gratin with quinoa |
|  | Snack: | 150g yoghourt (1.5% fat), 4 nuts |
|  | Supper: | 1 kiwi<br>Celery salad<br>Millet soufflé (normal soufflé with vegetable cubes with vegetable sauce and vegetables of the day)<br>Herbal tea or grain coffee |
| 4th food-reintroduction day<br>1600 kcal | Breakfast: | Muesli (quark, linseed oil, fruits, oats)<br>Quark-oil-cream and crispbread<br>Herbal tea and grain coffee |
|  | Lunch: | Quark-oil-cream<br>light dish of the day |
|  | Snack: | 20g nuts |
|  | Supper: | Raw carrot vegetables with apple<br>light dish of the day<br>Herbal tea and grain coffee |
| **Ketogenic food reintroduction** |  |  |
| Day of breaking the fast | Lunch: | 200g zucchini puree |
|  | 2:00 p.m. | 5 almonds |
|  | Supper: | 200g vegetables (e.g., 100g fennel + 100g carrots)<br>1 tbsp olive oil |
| 1st food-reintroduction day<br>800 kcal | Breakfast: | 100g dairy product mixed with 1 tbsp flaxseed oil<br>50g fruit<br>1 tbsp coconut flakes, 1 tbsp chia seeds<br>Herbal tea or grain coffee |
|  | Lunch: | Chicory salad with avocado vinaigrette<br>Quark-oil-cream<br>100g spinach and 100g broccoli<br>1 tbsp olive oil |
|  | Snack: | 5 Almonds |
|  | Supper: | 200ml vegetable soup (without potatoes)<br>200g vegetable plate (e.g., celery, tomato, kohlrabi)<br>1 tbsp vegan pesto, 1 tbsp olive oil<br>Herbal tea or grain coffee |
| 2nd food-reintroduction day<br>1000 kcal | Breakfast: | 150g dairy product mixed with 1 tbsp flaxseed oil<br>50g fruit<br>1 tbsp chopped almonds, 1 tbsp chia seeds<br>Herbal tea or grain coffee |
|  | Lunch: | Summer salad with green leaves, 100g tomato, 60g avocado, 2 tbsp sprouts, and 60ml avocado vinaigrette<br>Quark-oil-cream |
|  | Snack: | 20g walnuts |
|  | Supper: | Green salad with 1 grated fennel or kohlrabi, 1 tbsp pumpkin seeds, vinaigrette<br>Omelette made from 1 egg, served with 200g broccoli and 1 tbsp sunflower seeds<br>1 tbsp olive oil<br>Herbal tea or grain coffee |
| 3rd food-reintroduction day<br>1200 kcal | Breakfast: | 200g coconut yoghurt mixed with 1 tbsp flaxseed oil and 1 tbsp lemon juice<br>50g fruit<br>2 tbsp coconut flakes, 1 tbsp chia seeds<br>Herbal tea or grain coffee |
|  | Lunch: | Summer salad with green leaves, ½ pepper, 100g grated zucchini or radishes, 60ml avocado vinaigrette, 40g feta cheese, and 60g avocado<br>Quark-oil-cream<br>50g keto bread<br>1 tbsp olive oil |
|  | Snack: | 20g walnuts |
|  | Supper: | Green salad with 100g grated celery salad, 1 tbsp chopped almonds, and 60ml avocado vinaigrette<br>100g smoked tofu with 1 tsp olive oil<br>200g vegetable plate (e.g., 100g fennel, zucchini, spinach, or chard, with 1 tbsp pesto)<br>1 tbsp olive oil<br>Herbal tea or grain coffee |
| 4th food-reintroduction day<br>1600 kcal | Breakfast: | 200g dairy product mixed with 2 tbsp flaxseed oil and 1 tbsp lemon juice<br>50g fruit<br>1 tbsp chopped almonds, 1 tbsp sunflower seeds, 1 tbsp chia seeds<br>Herbal tea and grain coffee |
|  | Lunch: | Gazpacho (100g tomato, 200g cucumber, 100g bell pepper, 2 tbsp olive oil, and garnished with 1 tbsp chopped herbs, scallions, or cress)<br>Sautéed spinach with some onions in 1 tsp olive oil<br>Omelette (2 eggs with 1 tbsp Parmesan), served with 100g tomato sauce<br>1 tbsp olive oil |
|  | Snack: | 20g walnuts |
|  | Supper: | Mixed salad (green leaves, arugula, tomato, cucumber, zucchini, bell pepper, and herbs)<br>2 tbsp green olives |
|  |  | 3 tbsp olive oil and 2 tbsp lemon juice or vinegar dressing<br>100g goat cheese or 200g cottage cheese mixed with 1 tbsp<br>olive oil and herbs<br>Herbal tea and grain coffee |

**Table S3.** Baseline data comparison between the 32 closest neighbours of our participants extracted from subjects of our study with a cohort of 1422 participants. The distance computed was the Euclidean, based on the following normalised parameters: BMI (kg/m²), age, SBP, waist circumference, triglycerides. P-value is from a t-test.

|  | **MFP**<br>**(n =32)** | **Closest neighbours PF (n**<br>**=32)** | **p-value** |
| --- | --- | --- | --- |
| **Women (n, %)** | 22 (68.8) | 22 | 1 |
| **Men (n, %)** | 10 (31.2) | 10 | 1 |
| **Age (years)** | 41.5 ± 12.88 | 44.91 ± 12.76 | 0.292 |
| **BMI** | 25.35 ± 2.67 | 25.27 ± 2.61 | 0.897 |
| **Waist circumference** | 82.85 ± 9.44 | 84.69 ± 10.69 | 0.460 |
| **SBP** | 132.1 ± 15.4 | 127.84 ± 12.76 | 0.237 |
| **Triglycerides** | 88.44 ± 49.64 | 99.77 ± 44.76 | 0.341 |

**Table S4.**
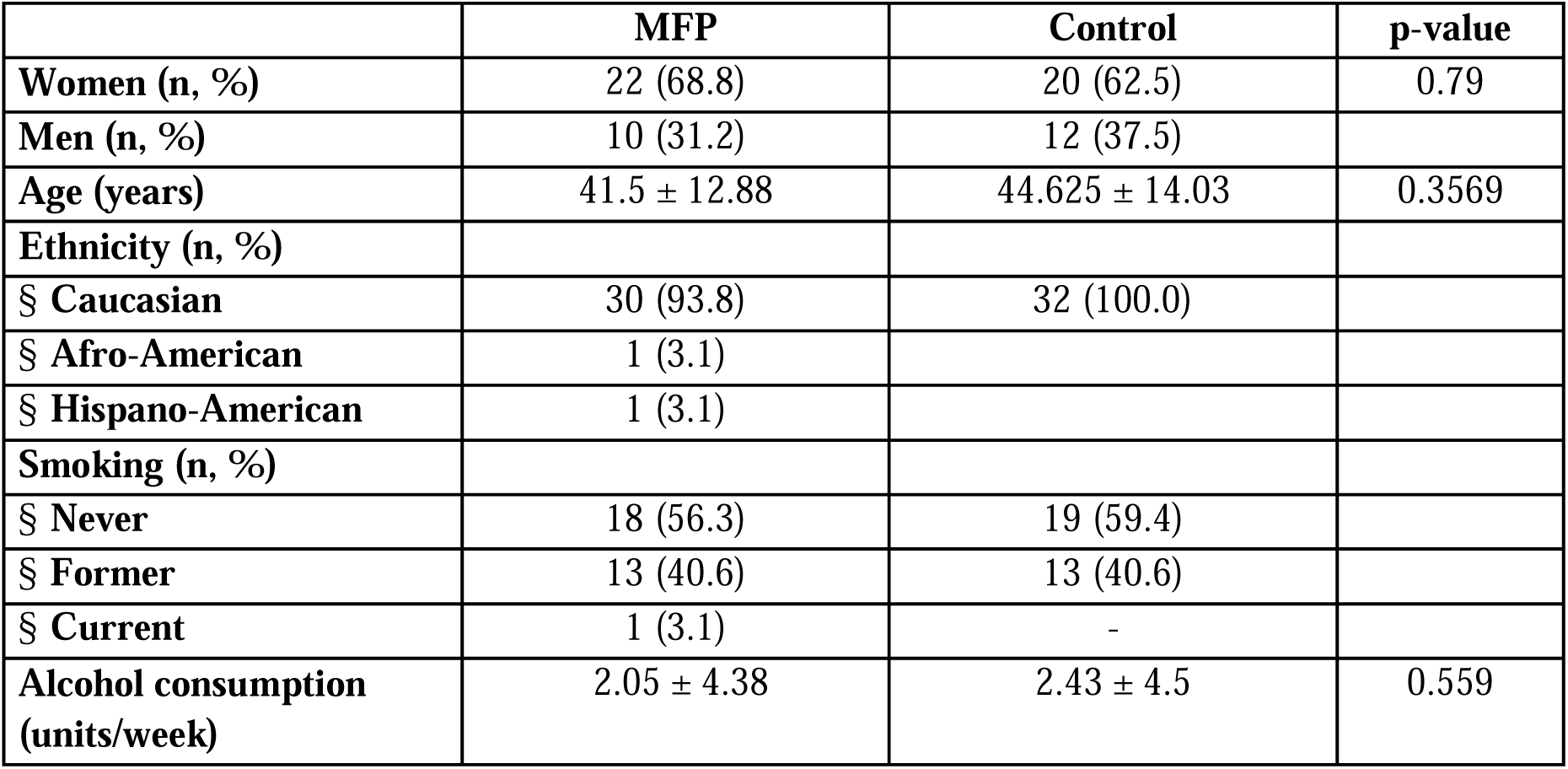
Study demographic data.

|  | **MFP** | **Control** | **p-value** |
| --- | --- | --- | --- |
| **Women (n, %)** | 22 (68.8) | 20 (62.5) | 0.79 |
| **Men (n, %)** | 10 (31.2) | 12 (37.5) |  |
| **Age (years)** | 41.5 ± 12.88 | 44.625 ± 14.03 | 0.3569 |
| **Ethnicity (n, %)** |  |  |  |
| § **Caucasian** | 30 (93.8) | 32 (100.0) |  |
| § **Afro-American** | 1 (3.1) |  |  |
| § **Hispano-American** | 1 (3.1) |  |  |
| **Smoking (n, %)** |  |  |  |
| § **Never** | 18 (56.3) | 19 (59.4) |  |
| § **Former** | 13 (40.6) | 13 (40.6) |  |
| § **Current** | 1 (3.1) | - |  |
| **Alcohol consumption**<br>**(units/week)** | 2.05 ± 4.38 | 2.43 ± 4.5 | 0.559 |

**Table S5.**
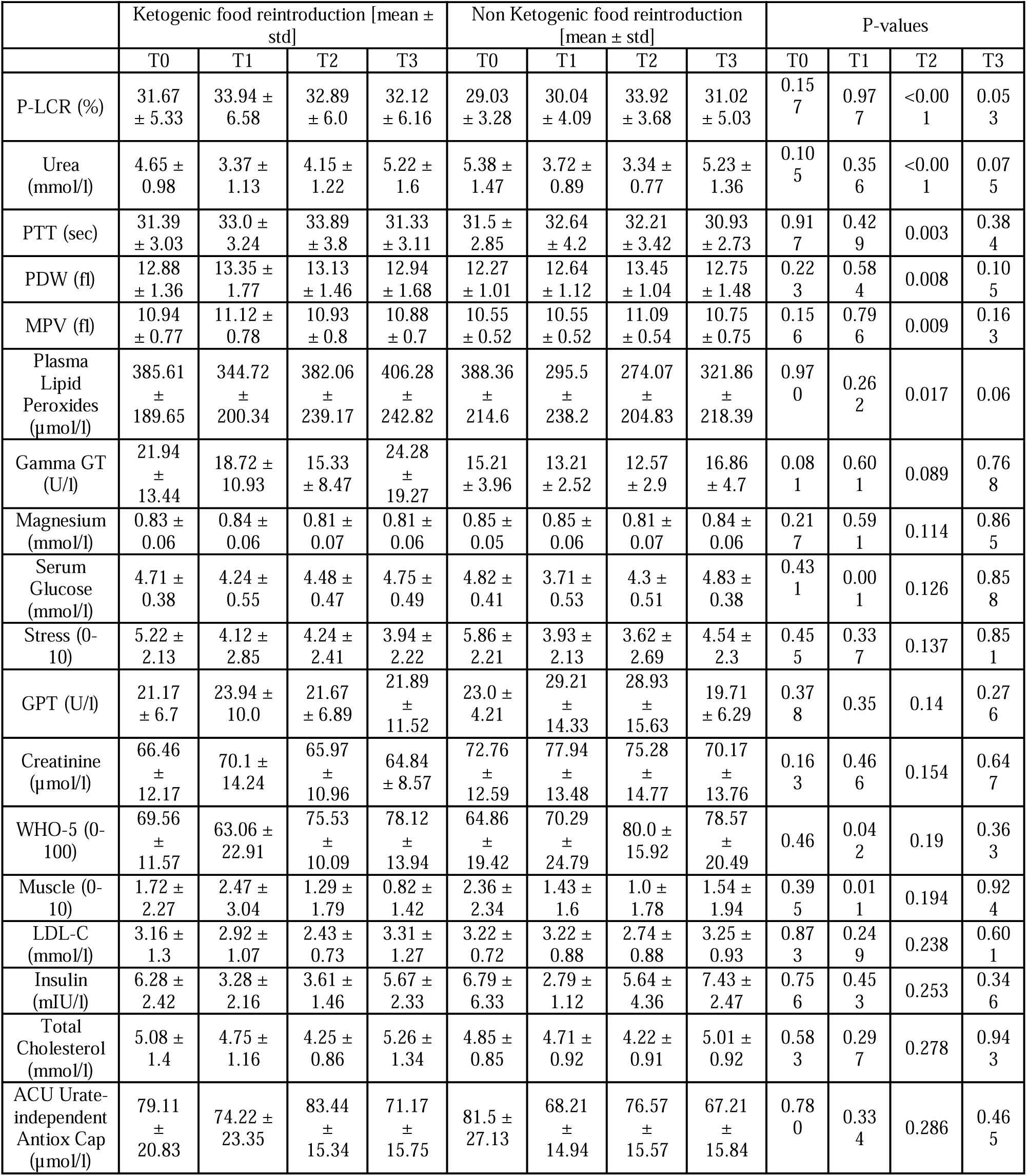
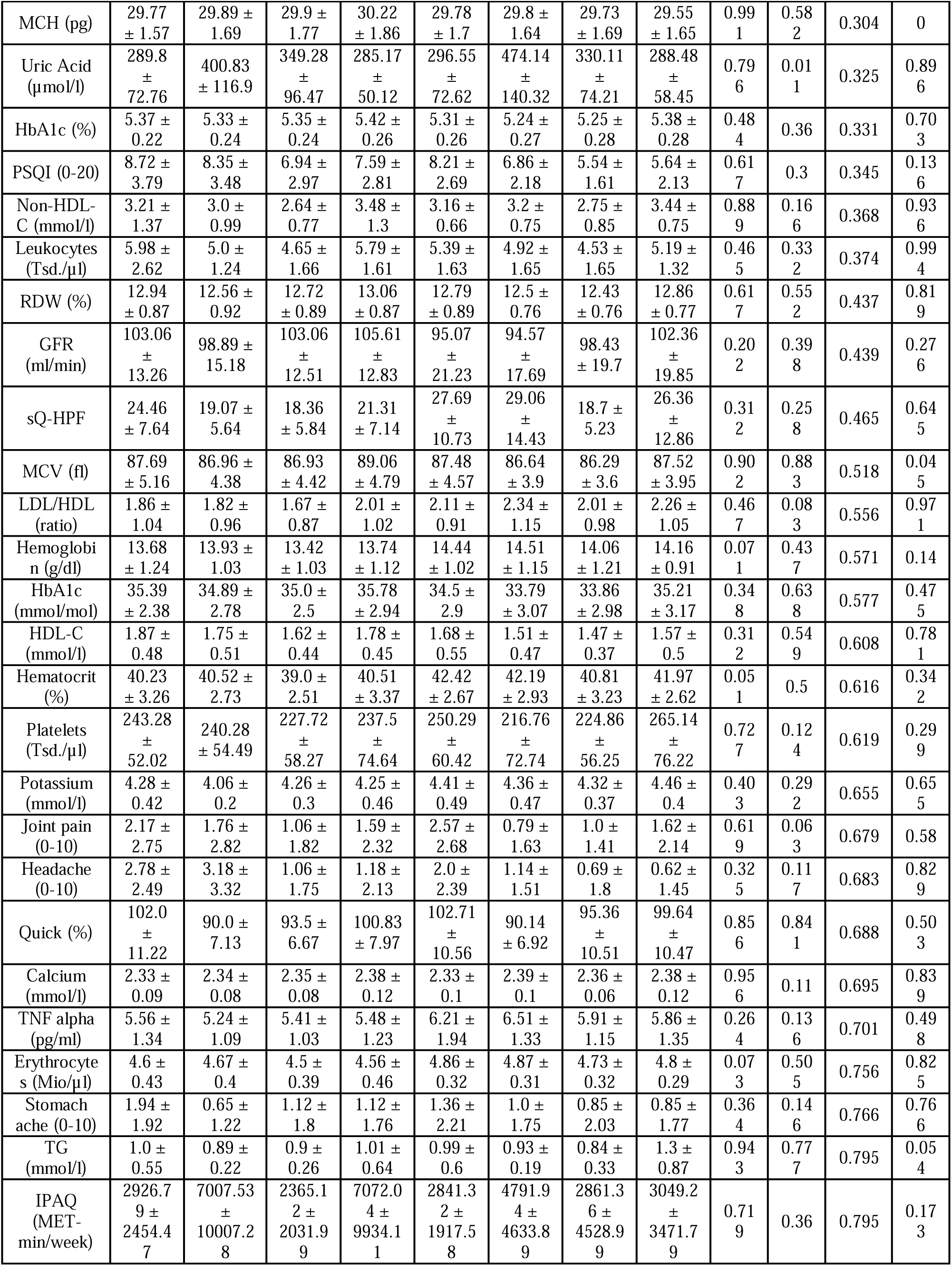
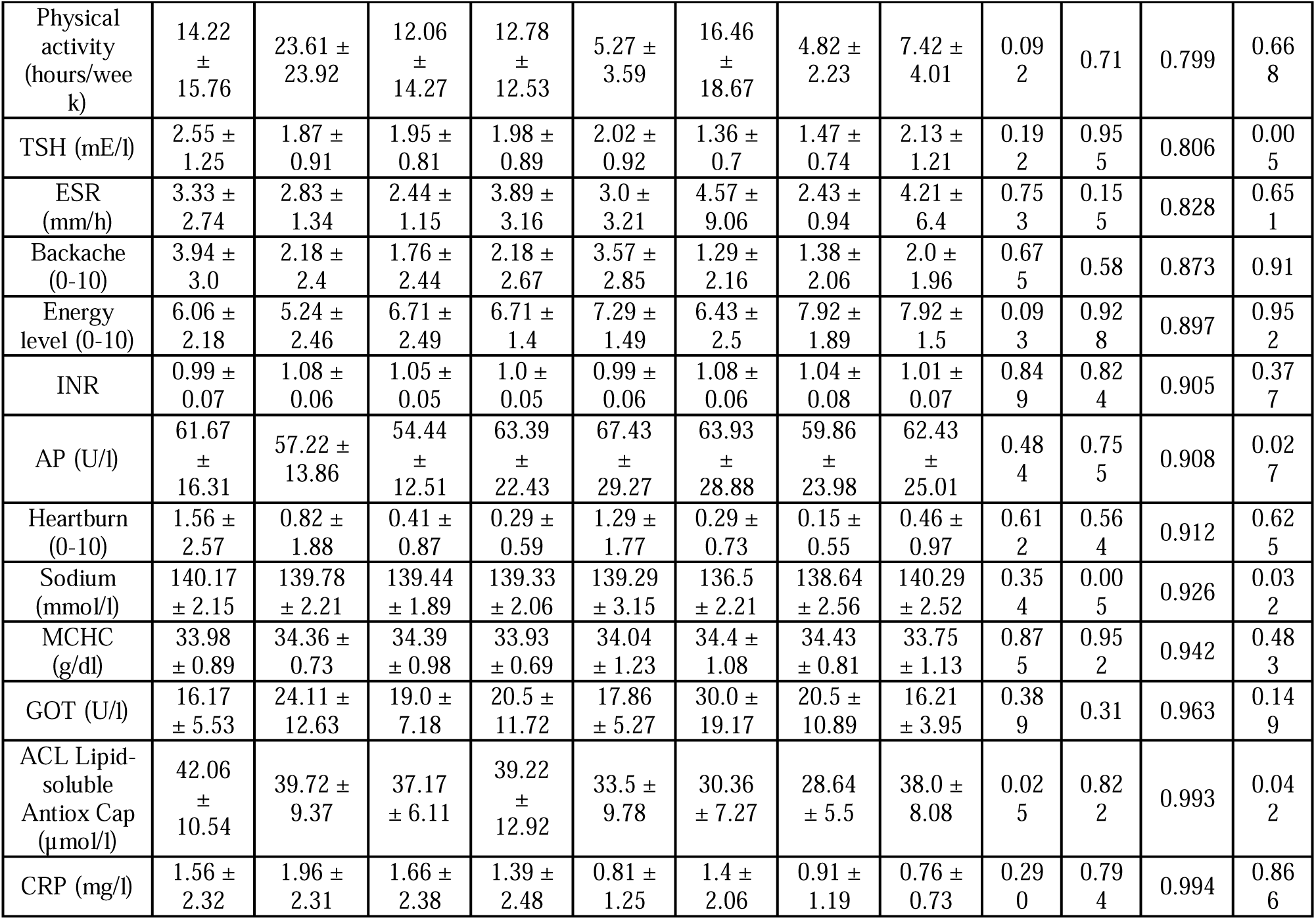
Differences in clinical parameters between the ketogenic food reintroduction and the standard food reintroduction. The p-value indicates the statistical significance in a linear-mixed model where the effects of the effects between the ketogenic food reintroduction and the non-ketogenic food reintroduction for the participants receiving the MFP at the same timepoint (*, p < 0.05 ; **, p < 0.01 ; ***, p < 0.001). # Primary endpoints.

**Table S6.**
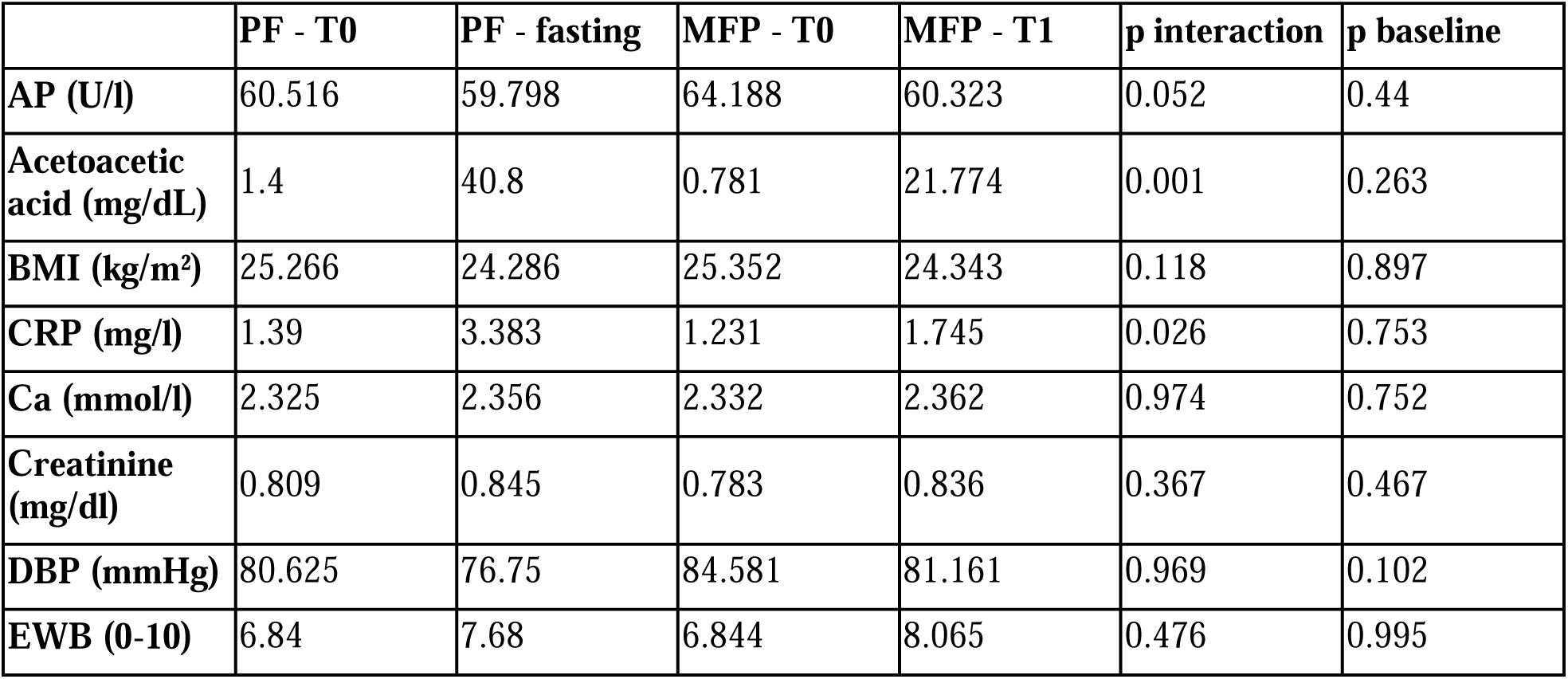
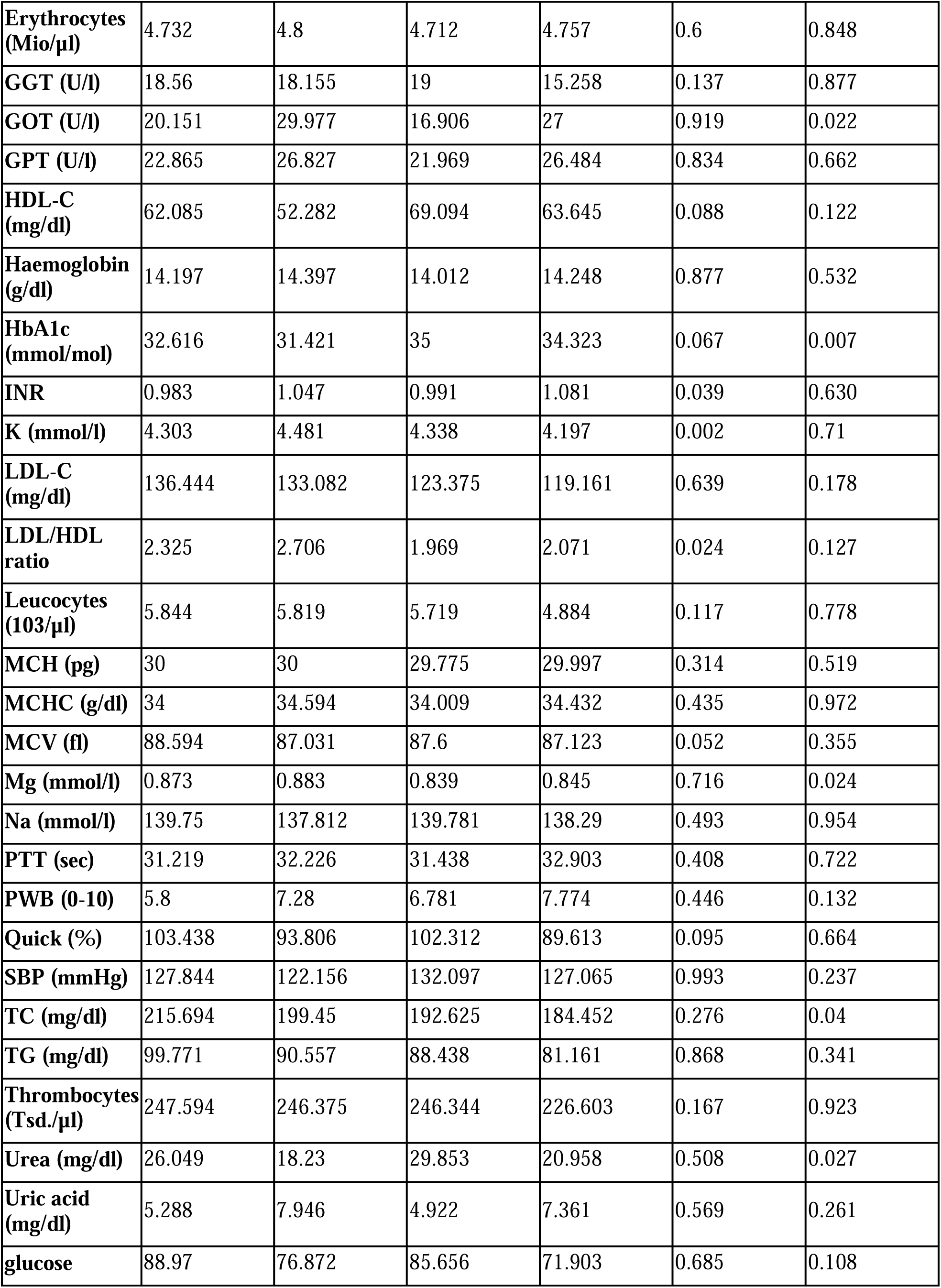
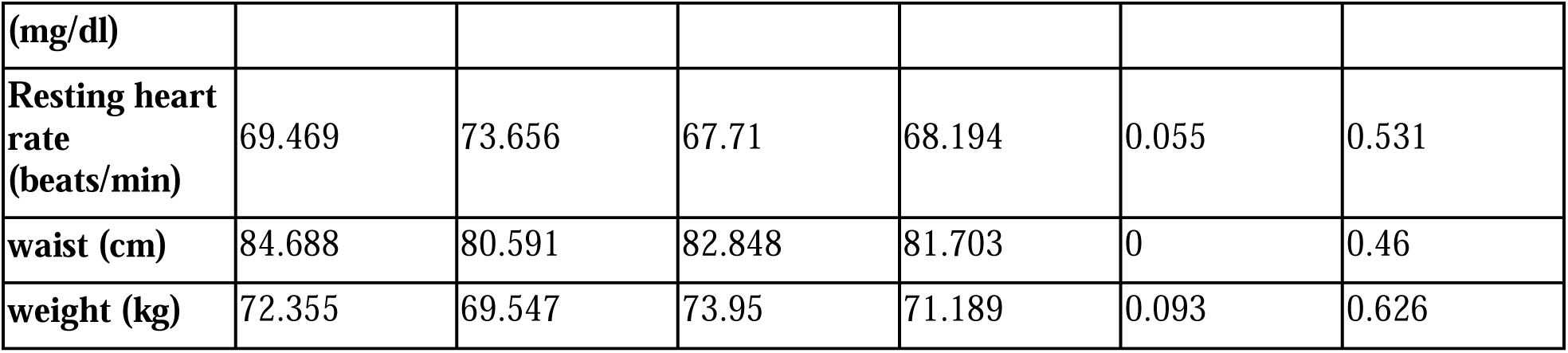
Comparison of the effects of the MFP and the PF. The effects of the MFP were compared to those of 32 matched individuals doing 5 day PF with a linear mixed model (participants as random effects). P interaction shows whether the changes between MFP and PF are different. The p baseline is the comparison of the MFP and PF baseline values.

